# Pitfalls and Solutions in Clone-Censor-Weight for Target Trial Emulation: Insights from Review, Simulation, and Real-World Analyses

**DOI:** 10.64898/2026.09.02.26362102

**Authors:** Yuya Kimura, Yuki Takazawa, Hideo Yasunaga

## Abstract

**Background:** The clone-censor-weight (CCW) method is increasingly finding application in target trial emulation to compare treatment strategies involving grace periods. However, its performance has not been systematically evaluated against the ground truth. Many studies utilising CCW ignore time-varying covariates and informative pre-existing censoring. Heavy-tailed inverse probability of censoring weights (IPCW) may yield biased estimates and under-coverage.

**Methods:** Simulation Part 1 evaluated the bias, root mean squared error, and coverage probability of CCW analysis under correctly specified and misspecified IPCW models across scenarios with or without time-varying covariates and informative pre-existing censoring. Part 2 varied the confounding strength, assessing whether inference failures could be detected using an IPCW tail-heaviness index (a weighted Hill estimator-derived Pareto-type tail index) and the estimator’s standard deviation, the target of bootstrap standard error.

**Results:** Under correct specification, all bias estimates were below 0.005; coverage approached the nominal level of 0.95 with increasing sample size. Omitting the time-varying covariate or mishandling pre-existing censoring yielded bias up to 0.069 and coverage substantially below 0.95. Under strong confounding, confidence intervals failed to attain nominal coverage, even with correct specification and large sample sizes, when the oracle tail-heaviness index was ≤1. When the index was >1, the coverage approached 0.95 as the estimator’s standard deviation decreased.

**Conclusions:** CCW analysis yields accurate estimates and confidence intervals with nominal coverage when implemented correctly, and confounding is not excessively strong. Investigators should account for time-varying covariates and pre-existing censoring and evaluate the tail behaviour of IPCW distribution and estimator’s bootstrap standard error.

**Key messages:**

– Correctly specified and implemented clone-censor-weight analyses can provide accurate strategy-specific risk estimates and confidence intervals with nominal coverage, whereas misspecified or incorrectly implemented analyses, such as those failing to account for time-varying confounders or informative pre-existing censoring, can yield biased estimates and under-coverage.
– Even with correct model specifications and large samples, strong confounding can lead to heavy-tailed distributions of the inverse probability of censoring weights and invalidate standard confidence interval construction.
– The tail heaviness index of the inverse probability of censoring weight distribution, in conjunction with the estimator’s bootstrap standard error, can help identify unreliable analyses and guide the specification of eligibility criteria and treatment strategies.

## Introduction

The use of target trial emulation (TTE) with real-world data, a framework mimicking randomised controlled trials (RCTs), has witnessed rapidly expansion. In TTE, investigators must align the timing of treatment assignment, eligibility assessment, and the commencement of outcome follow-up (time zero) [1]. However, treatment assignment is often not clearly determined at time zero. Therefore, investigators commonly introduce a grace period after time zero, assigning individuals to the intervention or control group according to whether they initiate treatment within this window. However, without appropriate adjustment, this approach can lead to immortal time bias. That is, individuals who would have initiated treatment during the grace period but experienced the outcome beforehand are misclassified into the control group. Landmark analysis mitigates this bias by restricting the analysis to individuals who remain event-free until the landmark is reached, thereby changing the target population from all eligible individuals at time zero to those who reach the landmark. The clone-censor-weight (CCW) method was proposed to overcome this problem [1,2]. This approach defines treatment strategies (e.g. initiating treatment within the grace period) and estimates the outcome risk under each predefined strategy. All eligible individuals are cloned and assigned to each strategy (the clone step), censored when they deviate from their assigned strategy (‘artificial censoring’, i.e. censor step), and weighted using inverse probability of censoring weighting (IPCW) to account for the resulting selection bias (weight step).

Despite its conceptual appeal, there are several important concerns regarding CCW analysis. First, no simulation study has systematically evaluated whether this approach yields accurate strategy-specific outcome estimates and confidence intervals with nominal coverage against a known ground truth. Notably, theoretical research on single-time-point inverse probability weighting has shown that the heaviness of the tail of the weight distribution governs the large-sample behaviour of the estimator and may compromise statistical inference [3]. This concern may be even more pronounced for IPCW because censoring weights accumulate over multiple time points, and artificial censoring may occur frequently. Second, almost half of the studies applying CCW analysis to real-world medical data did not account for time-varying covariates in their IPCW models, although such covariates generally affected deviations from the assigned strategy [4,5]. Third, most of these studies did not account for informative pre-existing censoring, such as loss to follow-up due to worsening clinical conditions, which must be appropriately addressed in both RCTs and TTE studies [5]. The details of our review of the second and third issues are provided in Supplementary File 1.

Given these concerns, this study had two objectives. First, we conducted simulation studies to examine whether CCW analysis can yield accurate strategy-specific outcome estimates and confidence intervals with nominal coverage when correctly modelled and implemented and confounding is not excessively strong, including scenarios with time-varying covariates affecting deviation from the assigned strategy and scenarios involving informative pre-existing censoring. Second, we examined whether confidence intervals could fail to attain nominal coverage under strong confounding, and consequently, heavier-tailed weight distributions, even when the CCW analysis was correctly implemented, and whether an IPCW tail-heaviness index could detect such failures.

## Methods

### General Principle of CCW Analysis

When implementing CCW analysis, investigators must first specify the target trial protocol, including the treatment strategies to be compared. The target estimand is the risk that would be observed if individuals fully adhere to their assigned treatment strategies.

The most basic treatment strategies in CCW analyses are: (1) initiating the treatment of interest within a pre-specified grace period (intervention strategy) and (2) not initiating it within the grace period (control strategy). The key considerations for specifying the treatment strategies and step-by-step description of the CCW approach are provided in Supplementary File 1.

### Simulation Study Part 1

Part 1 evaluated whether correctly implemented CCW analysis yielded accurate strategy-specific outcome estimates and confidence intervals with nominal coverage when confounding is not excessively strong.

### Data generation

The simulated datasets included six variables: three baseline covariates (age, sex, and Charlson Comorbidity Index) [6], one time-varying covariate [blood oxygen saturation (SpO_2_)], treatment, and the outcome of in-hospital death. In Scenario 3, an additional variable indicating pre-existing censoring was generated. In these scenarios, patients were assumed to be hospitalised for an acute respiratory infection; the baseline covariate distributions, longitudinal dynamics of SpO_2_, and their associations with treatment, outcome, and pre-existing censoring were specified accordingly. The directed acyclic graph (DAG) describing the causal relationship of the simulated data, together with the variable definitions and specified effects, is provided in Supplementary Files 1 and 2. We considered three scenarios that differed according to the presence or absence of a time-varying covariate affecting treatment decisions and the outcomes and informative pre-existing censoring. Using 1 and 0 to denote presence and absence, respectively, the distribution of (time-varying covariate, informative pre-existing censoring) was (1, 0) in Scenario 1, (0, 0) in Scenario 2, and (1, 1) in Scenario 3.

For the ground truth, we generated 10 000 000 individuals per scenario, all forced to adhere to each predefined treatment strategy (Supplementary File 1), to estimate the potential outcome under full adherence. For the CCW simulations, we generated 1000 datasets per scenario and sample-size setting (100, 1000, and 10 000) according to the specified DAG.

### Evaluation metrics

The 30-day outcome risk under each treatment strategy was estimated using an IPCW Kaplan–Meier estimator, with 95% confidence intervals obtained by bootstrap resampling with 1000 replicates. Performance was evaluated across 1000 simulated datasets using bias (difference between the average point estimate and simulated ground-truth 30-day outcome risk), average root mean squared error (RMSE), and coverage probability (the proportion of datasets in which the bootstrap 95% confidence interval contained the ground truth value).

### Experimental overview

For each scenario, we conducted experiments to compare the correctly specified and misspecified models (Table 1). The intervention and control strategies were defined as ‘initiating’ and ‘not initiating treatment within the grace period’, respectively, with the last (cut-off) day of the grace period set to day 0 (day of admission), 2, or 4. Experiments A and B used data generated in Scenario 1, and did and did not include the time-varying covariates in the IPCW model, respectively. Experiment C used Scenario 2, which included only baseline covariates. Experiments D–F used Scenario 3. Experiment D fitted separate IPCW models for artificial and pre-existing censoring and multiplied the resulting weights. Experiment E fitted a single IPCW model using a composite censoring indicator. Experiment F did not account for pre-existing censoring. Accordingly, Experiments A, C, and D at all cut-off days, together with Experiment B at cut-off day 0 (where no time-varying covariate existed), represented the correctly specified models, whereas Experiment B at cut-off days 2 and 4, and Experiments E and F represented misspecified settings.

### Simulation Study Part 2

Part 2 evaluated whether confidence intervals could fail to attain nominal coverage in settings with strong confounding and whether an IPCW tail-heaviness index could detect such failures.

### Data generation

Using Scenario 1 from Part 1 as the reference, we generated datasets with different confounding strengths by multiplying the covariate effects on either treatment assignment or outcome occurrence by 0.5, 2, 4, or 8.

### Evaluation metrics

The 30-day outcome risk and 95% confidence interval for each treatment strategy were estimated using the same methods as in Part 1. Part 2 focused on the coverage probability because all models were correctly specified; therefore, bias and RMSE were not the primary evaluation metrics.

### Tail-heaviness index for IPCW

To assess sensitivity to extreme IPCW components, we used a heuristic tail-heaviness diagnostic motivated by the variance decomposition of the IPCW hazard score and previous tail-index analyses of IPW estimators [3]. At each grace period time point, among individuals still at risk, we applied a weighted version of the Hill estimator [7] to the inverse estimated probability of remaining uncensored, using the squared inverse cumulative uncensoring probability up to the previous time point as weights. Smaller indices indicated heavier tails. Under a Pareto-type approximation [8] and finite-mean historical component, a population index below 1 implies an infinite second moment of the hazard score. However, estimated values below 1 should be viewed as warnings rather than proof of infinite variance. We calculated the indices by strategy and time point, using the minimum as a summary. We also obtained oracle benchmark estimates from 1 000 000 individuals using true censoring probabilities. Intuitive explanations and further details are provided in Supplementary Files 1 and 2, respectively.

### Experimental overview

Because Part 2 presupposed the correct model specification, we limited it to Experiment A; the treatment strategies and grace period settings were identical to those in Part 1.

First, at a sample size of 10 000, we assessed whether the coverage probability fell below the nominal level of 0.95 across different levels of confounding strength and whether such failures could be detected using the oracle tail-heaviness index. Second, we examined how coverage varied across sample sizes of 100, 1000, and 10 000, stratified by whether the oracle index was >1 or ≤1, hypothesising that coverage would approach 0.95 with increasing sample size when the index was >1. Third, we calculated the standard deviation of the estimated outcome risk across 1000 Monte Carlo replications—the quantity targeted by the bootstrap standard error, a practically obtainable indicator of whether the sample size is sufficient in non-simulation settings—and examined its relationship with empirical coverage. Fourth, we assessed the discrepancy between the oracle and estimated tail-heaviness indices.

Additional explanations for the simulation studies are provided in Supplementary File 2; the corresponding code is available in our GitHub repository (https://github.com/yukiregista/clone-censor-weight.git).

### CCW Analysis Using Real-World Medical Data

To illustrate the key components that should be reported in a CCW analysis and demonstrate the differences between analyses that did and did not account for informative pre-existing censoring, we emulated a target trial evaluating whether bronchial artery embolisation, a catheter-based treatment performed within 5 days of hospitalisation, reduced in-hospital mortality among patients admitted with severe haemoptysis (Supplementary File 1).

## Results

### Simulation Study Part 1

The patients’ characteristics and observed and estimated 30-day outcome risks in Scenarios 1, 2, and 3 are presented in Supplementary File 1. In Experiments A, B (cut-off day 0), C, and D, in which the models were correctly specified, all bias estimates were below 0.005 (Fig. 1). In contrast, the bias reached 0.025 in Experiment B, 0.069 in Experiment E, and 0.028 in Experiment F.

**Figure 1.**
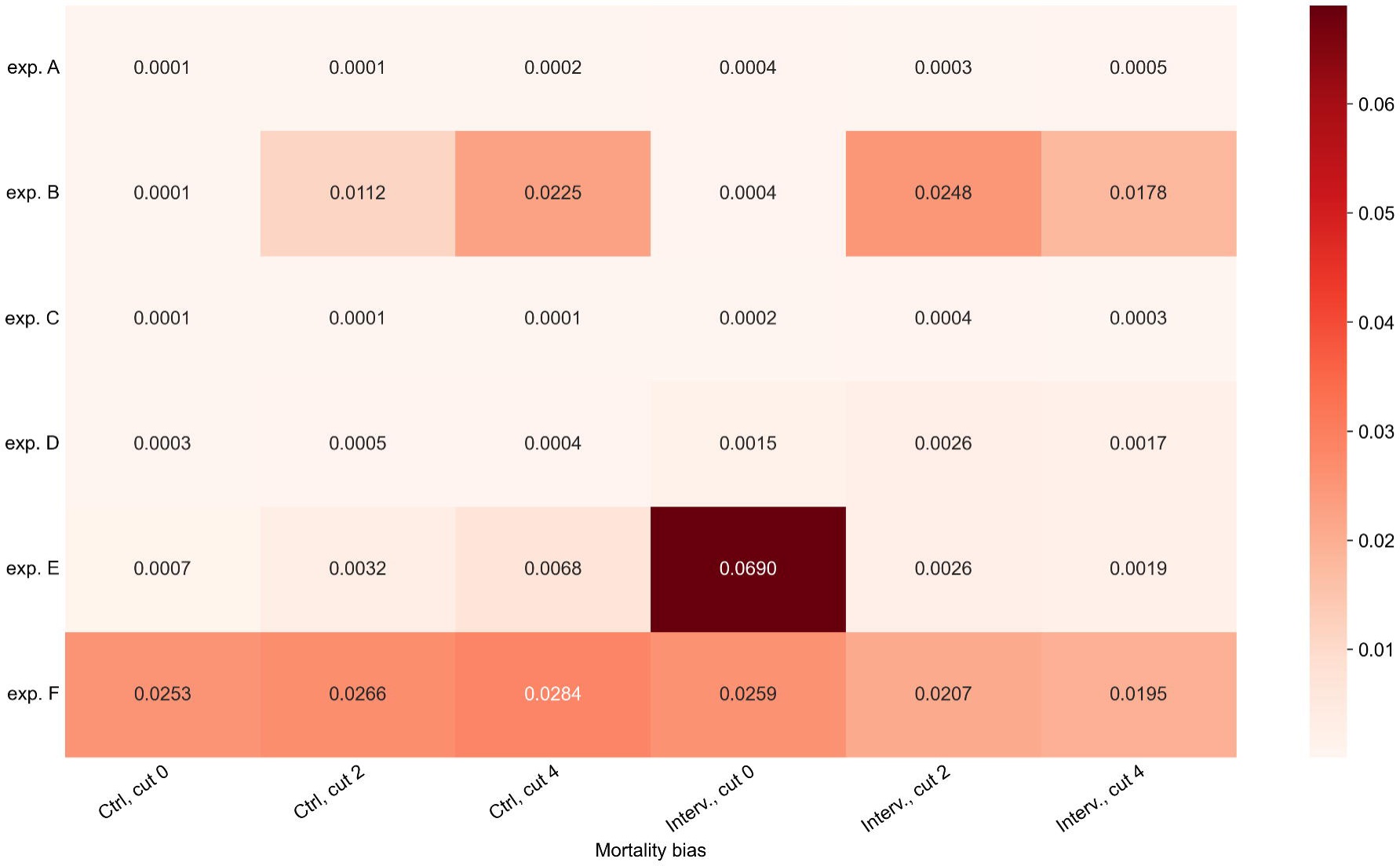
Bias in clone-censor-weight estimates relative to the ground truth simulation. Experiments A, C, and D represent correctly specified analyses for their respective scenarios, whereas Experiments B, E, and F represent misspecified alternatives: omission of time-varying covariates (B), joint rather than separate modelling of artificial and pre-existing censoring (E), and no adjustment for pre-existing censoring (F). The results are based on experiments with a sample size of 10 000. Alt text: Plot of bias against the ground-truth 30-day risk for Experiments A to F, shown separately for the control and intervention strategies at cut-off days 0, 2 and 4, with a sample size of 10 000. Bias is below 0.005 for Experiments A, C, and D and for Experiment B at cut-off day 0, and increased to 0.069 for Experiment E, 0.028 for Experiment F, and 0.025 for Experiment B at cut-off days 2 and 4.

Correctly specified models generally performed better with respect to the RMSE than alternative specifications, except Experiment E that yielded a lower RMSE in some settings on cut-off days 2 and 4 (Fig. 2). In Scenario 1, Experiment B consistently had higher RMSE values compared with Experiment A at cut-off days 2 and 4. In Scenario 3, Experiment F generally had the highest RMSE, except for the intervention strategy at cut-off day 0, where Experiment E had the highest RMSE.

**Figure 2.**
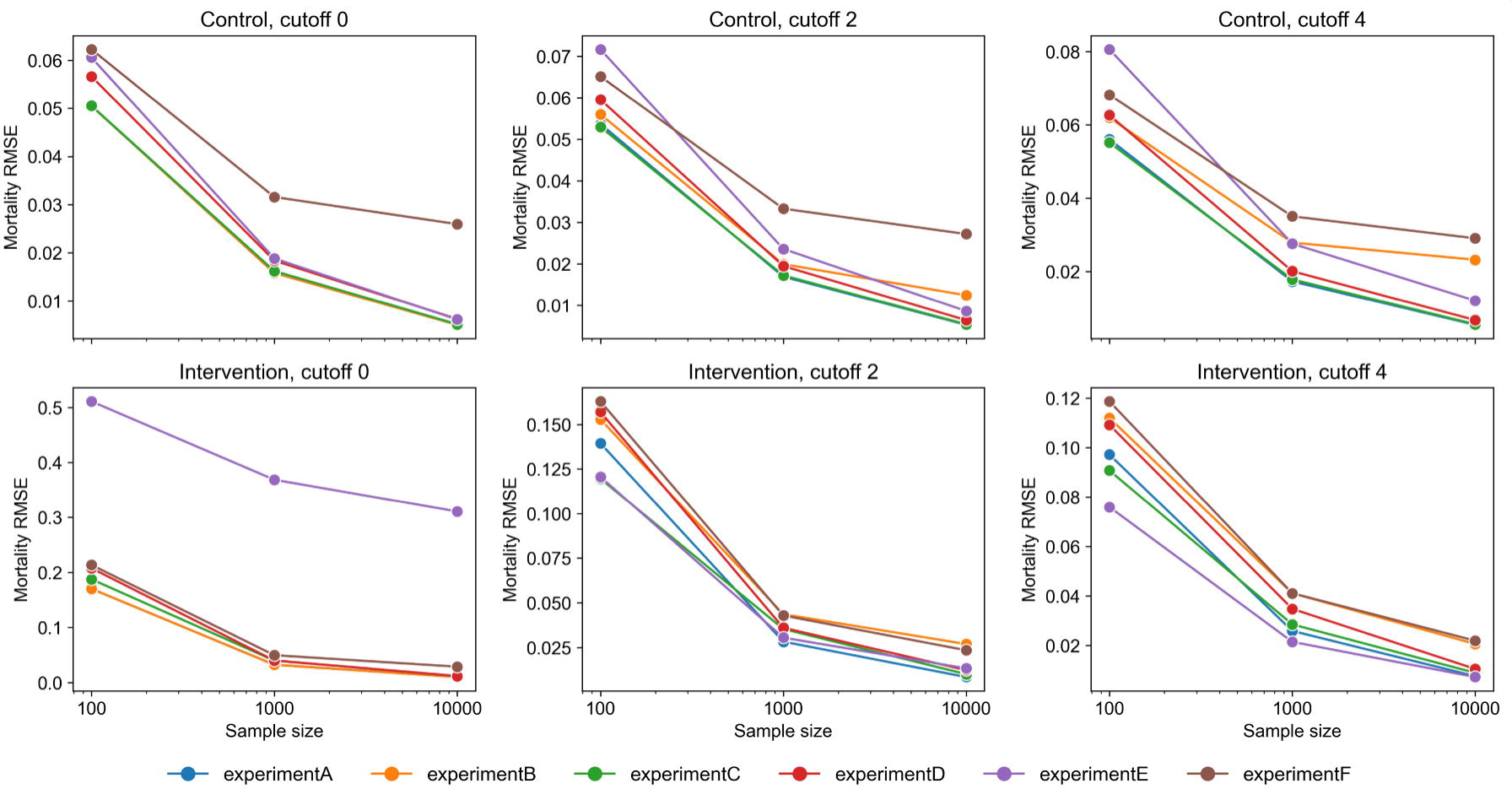
Root mean squared error of clone-censor-weight estimates relative to the ground truth simulation. Experiments A, C, and D represent correctly specified analyses for their respective scenarios, whereas Experiments B, E, and F represent misspecified alternatives: omission of time-varying covariates (B), joint rather than separate modelling of artificial and pre-existing censoring (E), and no adjustment for pre-existing censoring (F). The results are stratified by sample size (100, 1000, and 10 000). Alt text: Plot of root mean squared error for Experiments A to F, shown separately for the control and intervention strategies at cut-off days 0, 2, and 4, and stratified by sample sizes of 100, 1000 and 10 000. The root mean squared error decreases as the sample size increases and is generally the lowest for the correctly specified experiments.

Correctly specified models achieved coverage close to 0.95 for the control strategy across sample sizes and approached 0.95 for the intervention strategy at a sample size of 10 000, whereas misspecified models showed substantial under-coverage, often worsening as the sample size increased (Fig. 3).

**Figure 3.**
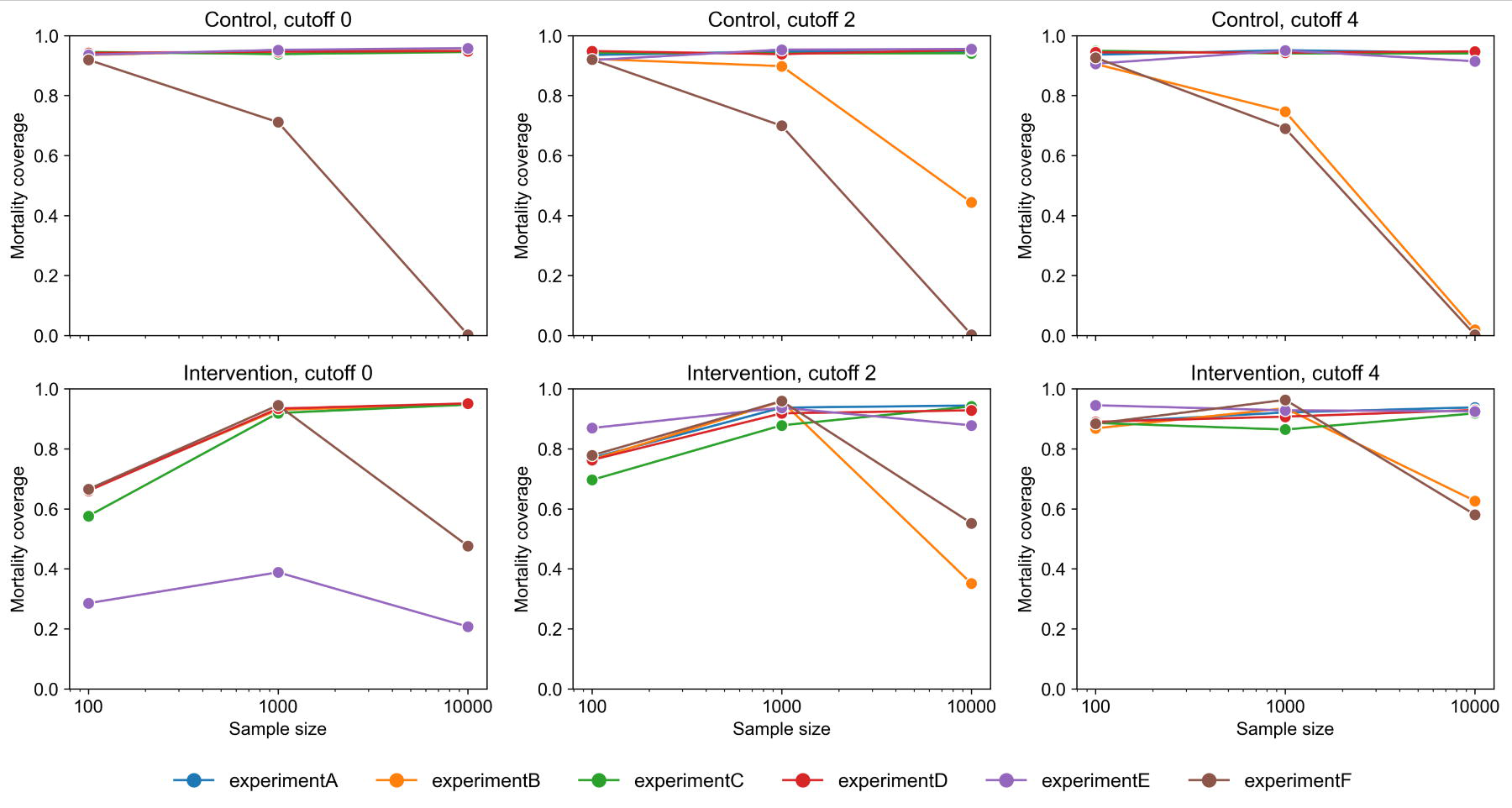
Coverage probability of clone-censor-weight estimates relative to the ground truth simulation. Experiments A, C, and D represent correctly specified analyses for their respective scenarios, whereas Experiments B, E, and F represent misspecified alternatives: omission of time-varying covariates (B), joint rather than separate modelling of artificial and pre-existing censoring (E), and no adjustment for pre-existing censoring (F). The results are stratified by sample size (100, 1000, and 10 000). Alt text: Plot of coverage probability of the bootstrap 95% confidence intervals for Experiments A to F, shown separately for the control and intervention strategies at cut-off days 0, 2, and 4, and stratified by sample sizes of 100, 1000 and 10 000, with a reference line at the nominal level of 0.95. Correctly specified experiments lie close to 0.95 for the control strategy at all sample sizes and approach 0.95 for the intervention strategy at a sample size of 10 000, whereas misspecified experiments fall well below 0.95, often further as the sample size increases.

### Simulation Study Part 2

The patients’ characteristics and observed and estimated 30-day outcome risks are presented in Supplementary File 1.

Figure 4 presents the results obtained when the strengths of the associations between the covariates and treatment decisions varied. At a sample size of 10 000, all settings with an oracle tail-heaviness index of >1 achieved coverage approximating the nominal level of 0.95 for both strategies, whereas those with an index ≤1 did not [panels (a) and (b)]. Coverage increased with sample size and approached 0.95 in settings with an index >1 but remained below the nominal level when the index was ≤1 [panel (c)]. In settings with an index >1, the coverage increased as the estimator’s standard deviation decreased. Nominal coverage was not achieved with a standard deviation of ≥ 0.10, was achieved in some settings at approximately 0.05, and in all at approximately 0.01 [panel (d)]. The estimated tail heaviness indices were generally similar to the corresponding oracle indices for both types of confounding changes (Supplementary File 1).

**Figure 4.**
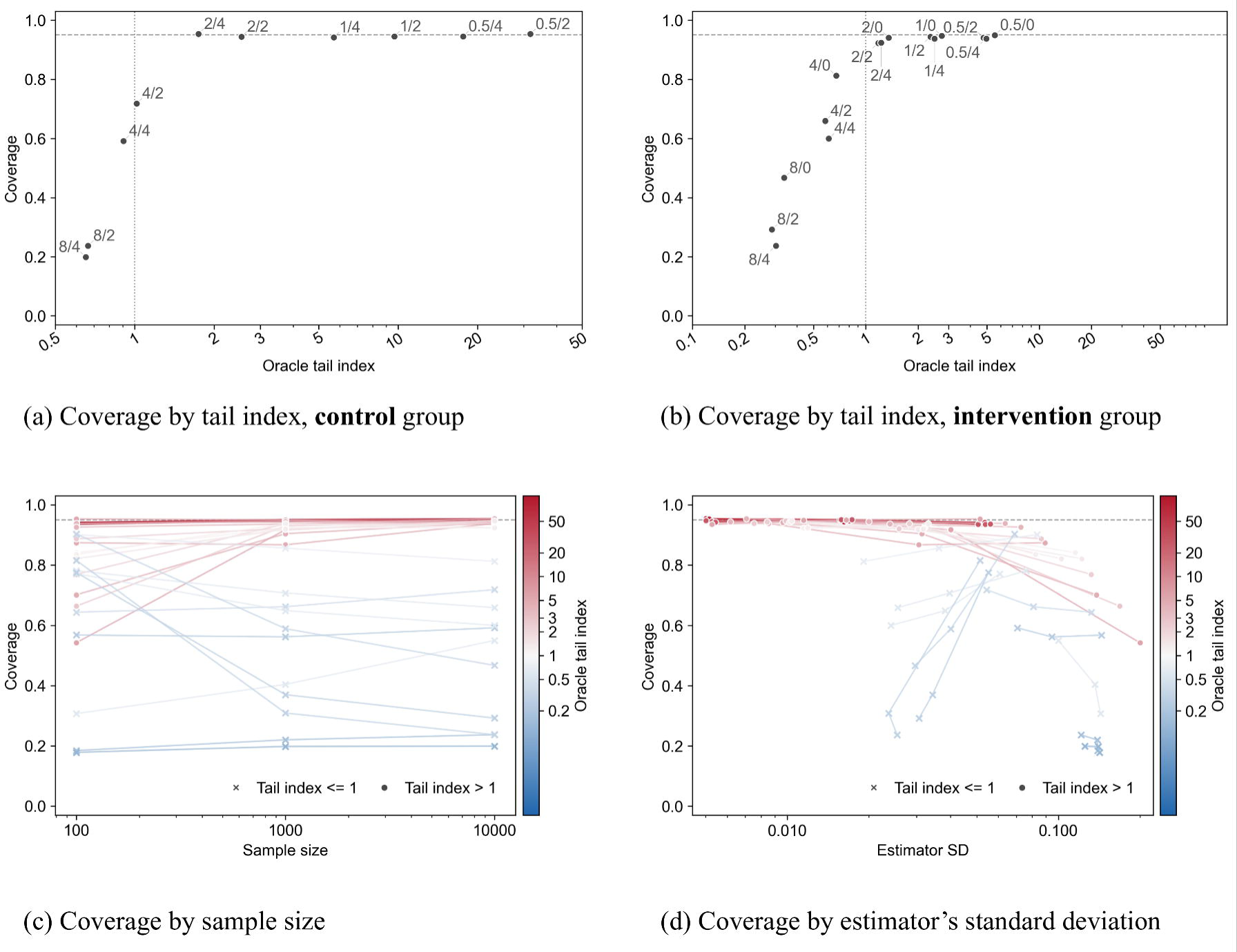
Coverage probability according to the oracle tail-heaviness index for inverse probability of censoring weights, sample size, and standard deviation of the estimator under varying strengths of confounding affecting treatment assignment. This figure shows the coverage probability when the magnitude of the covariate effects on treatment assignment was varied. Panels (a) and (b) show the relationship between the oracle tail-heaviness index for inverse probability of censoring weights (IPCW) and coverage probability under the control and intervention strategies, respectively. In labels of the form “x/y” (e.g., 4/2), x denotes the factor by which the covariate effects on treatment assignment were multiplied relative to the reference setting used in Part 1, and y denotes the cut-off day, defined as the final day of the grace period. Panel (c) shows the coverage probability across sample sizes stratified by the oracle IPCW tail heaviness index. Panel (d) shows the relationship between the standard deviation of the estimator and coverage probability stratified by the oracle IPCW tail-heaviness index. Alt text: Four-panel figure on coverage probability when the magnitude of the covariate effects on treatment assignment is varied. Panels (a) and (b) plot the coverage probability against the oracle tail-heaviness index of the inverse probability of censoring weights for the control and intervention strategies, respectively, with settings labelled by the multiplier and cut-off day; settings with an index above 1 lie close to the nominal level of 0.95, and those with an index of 1 or below lie below it. Panel (c) plots the coverage probability across sample sizes of 100, 1000 and 10 000, stratified by whether the oracle index is above 1 or not. Panel (d) plots the coverage probability against the standard deviation of the estimator, stratified in the same way, showing coverage approaching 0.95, as the standard deviation falls towards 0.01.

The results of the simulations in which the magnitude of the covariate effects on the outcome varied are presented in Supplementary file 1.

### CCW Analysis Using Real-World Medical Data

Accounting for live discharge as pre-existing censoring, the 30-day all-cause in-hospital mortality was 28.7% (95% confidence interval, 25.9 to 31.2%) under the intervention strategy and 37.5% (35.4 to 39.5%) under the control strategy. The corresponding IPCW tail-heaviness indices were 12.7 and 8.5, with bootstrap standard errors of 0.010 and 0.014. In exploratory analyses ignoring pre-existing censoring, mortality was higher under both strategies (Supplementary File 1).

## Discussion

Our simulation studies yielded two main findings. First, Simulation Study Part 1 showed that CCW analysis can yield accurate strategy-specific outcome estimates and confidence intervals with nominal coverage when correctly modelled and implemented, and when confounding is not excessively strong, whereas performance deteriorated under misspecification. Second, Simulation Study Part 2 showed that attaining confidence intervals with nominal coverage can fail even with correct implementation and a large sample size, if strong confounding yields a heavy-tailed IPCW distribution. Coverage approached 0.95 with an increase in the sample size when the oracle IPCW tail-heaviness index was >1, but not when the index was ≤1.

Time-varying covariates that affect deviation from the assigned strategy must be accounted for when estimating the risk and causal effect under full adherence in RCTs and TTEs [4,5]. Nevertheless, approximately half of the real-world medical studies applying CCW analysis did not incorporate time-varying covariates, and our simulations demonstrated that omitting such covariates can cause under-coverage. Investigators should use clinical knowledge, prior evidence, and the assumed causal structure to pre-specify time-varying covariates associated with both artificial censoring and the outcome, incorporate them into the IPCW model, and justify any omissions.

Informative pre-existing censoring must also be addressed when estimating the risk and causal effect under full adherence [5]; however, most studies applying CCW analysis to real-world medical data failed to account for, or even mention, it. When informative pre-existing censoring is adjusted for, the resulting estimand represents the effect under a hypothetical condition in which neither strategy deviation nor such censoring occurs. Artificial censoring results from strategy deviation, whereas pre-existing censoring (e.g. loss to follow-up, live discharge, or loss of insurance eligibility) generally arises through different mechanisms; therefore, the covariates associated with the two processes and the magnitudes of their effects are unlikely to be identical. Indeed, Experiment D, which fitted separate IPCW models for the two censoring processes and multiplied the resulting weights, yielded low bias and near-nominal confidence interval coverage. Therefore, these distinct mechanisms should be reflected appropriately in the modelling approach, e.g. by fitting separate IPCW models.

Although we did not examine competing events, their statistical handling is technically similar to that of pre-existing censoring. However, the decision to treat a competing event as censoring and adjust for it via IPCW is not merely a matter of model specification, but a choice of estimand [9,10]. Censoring a competing event conceptually corresponds to estimating the outcome under a hypothetical condition in which the competing event does not occur. When an intervention that would eliminate the competing event is not categorically defined, the interpretation of this estimand may be unclear. Investigators should select an approach consistent with the study objective, such as a hypothetical composite treatment policy or competing risk strategy [11].

Nevertheless, Experiment E yielded lower RMSE values in some settings with a smaller empirical standard error (e.g. at cut-off 4 and n=10 000, the empirical standard errors for the intervention arm were 0.010 and 0.007 for experiments D and E, respectively; full results not shown). This may reflect implicit weight stabilisation from modelling the two censoring processes jointly, which reduced the occurrence of extreme weights. The potentially counterintuitive findings of Simulation Study Part 1 are further explained in Supplementary File 1.

The findings of Simulation Study Part 2 suggest that the practical application of CCW analysis entails evaluation of the tail behaviour of IPCW distribution. Because the estimated tail-heaviness indices were generally similar to their oracle values, an estimated index >1 may indicate that a valid large-sample inference is possible. Even when the index is >1, investigators should assess whether the available sample size is sufficient for which the bootstrap standard error of the estimator may provide a practical indicator. A possible diagnostic procedure is as follows. First, the IPCW tail-heaviness index should be estimated from the observed dataset. If it is smaller than or close to 1, investigators should reconsider the eligibility criteria or treatment strategies to avoid target populations containing many individuals with near-zero probabilities of adhering to a given strategy, thereby improving the tail behaviour of the weights. Second, if the index is >1, investigators should calculate the estimator’s bootstrap standard error and confirm that it is not excessively large. In our simulations, all examined settings with an estimator standard deviation of approximately 0.01 achieved nominal coverage. These values should be interpreted as empirical reference points rather than universal thresholds.

We also conducted a CCW analysis using real-world medical data to illustrate how the method can be implemented and reported; this analysis is discussed in Supplementary File 1.

As the application of TTE continues to expand, CCW analysis is likely to be increasingly applied to treatment strategies involving grace periods. Valid application requires careful consideration of the treatment strategy specification, artificial censoring, IPCW model specification, pre-existing censoring, competing events, and distribution of the resulting weights. To help investigators assess these elements systematically, we have provided a checklist for the design, implementation, and reporting of CCW analyses in Supplementary File 4.

This study had some limitations. First, although our simulation scenarios and experiments were designed to reflect realistic applications, we did not examine all possible data-generating mechanisms, censoring mechanisms, or forms of model misspecifications. Second, to maintain clarity and limit the computational burden, we restricted the number of covariates included in the simulations and the range of models evaluated and did not examine weight stabilisation or truncation, whose finite-sample bias–variance trade-offs warrant further investigation.

## Ethics approval

This study was approved by the Institutional Review Board of the University of Tokyo (approval number: 3501– (5)) and conducted in accordance with the Declaration of Helsinki. Given the retrospective use of de-identified medical data (extracted from the Diagnosis Procedure Combination database), the requirement for written informed consent was formally waived.

## Acknowledgements

In this research, we used UTokyo Azure (https://utelecon.adm.u-tokyo.ac.jp/en/research_computing/utokyo_azure/) for conducting simulation studies.

## Author contributions

Y.K.: conceptualisation, methodology, formal analysis, investigation, data curation, visualisation, project administration, writing—original draft, review, and editing. Y.T.: conceptualisation, methodology, formal analysis, investigation, data curation, visualisation, project administration, writing—review, and editing. H.Y.: Funding acquisition, supervision, writing, review, and editing.

Y. K. is the guarantor and takes responsibility for the integrity of the work

## Supplementary data

Supplementary data are available at IJE online.

## Data availability

The simulated data are available from our GitHub repository (https://github.com/yukiregista/clone-censor-weight.git). The real-world medical data analysed in this study (Diagnosis Procedure Combination database) are not publicly available because of data use agreements with the participating hospitals and data providers.

## Use of artificial intelligence (AI) tools

The authors used AI tools, including Claude (Fable 5 and Opus 5) and ChatGPT (GPT-5.6), to refine the language of the English manuscript and to assist with code generation and code revision for the simulation study. All AI-generated outputs were reviewed, verified, and edited by the authors, who take full responsibility for the final content of the manuscript.

