## Supplementary material for "Pitfalls and Solutions in Clone-Censor-Weight for Target Trial Emulation: Insights from Review, Simulation, and Real-World Analyses": Table

Tables

**Table 1. Summary of experiments in Simulation Study Part 1**

| **Scenario ID** | **Experiment ID** | **Include time-varying covariates in the IPCW model?** | **Account for pre-existing informative censoring?** | **Model pre-existing and artificial censoring separately?** | **Time terms in the IPCW model (control arm)?** | **Time terms in the IPCW model (intervention arm)?** |
| --- | --- | --- | --- | --- | --- | --- |
| 1^a^ | A | Yes | –^b^ | –^b^ | Yes^d^ | No^e^ |
| 1^a^ | B | No | –^b^ | –^b^ | Yes^d^ | No^e^ |
| 2^a^ | C | No | –^b^ | –^b^ | Yes^d^ | No^e^ |
| 3^a^ | D | Yes | Yes | Yes | Yes^d^,^f^ | No^e^,^f^ |
| 3^a^ | E | Yes | Yes | No | No^g^ | No^g^ |
| 3^a^ | F | Yes | No | –^c^ | Yes^d^ | No^e^ |

Three scenarios were defined based on the presence or absence of time-varying covariates and informative pre-existing censoring. Experiments A, C, and D represent correctly specified analyses for their respective scenarios, whereas Experiments B, E, and F represent misspecified alternatives: omission of time-varying covariates (B), joint rather than separate modelling of artificial and pre-existing censoring (E), and no adjustment for pre-existing censoring (F).

a. The pair (time-varying covariate, pre-existing informative censoring) is coded as follows: Scenario 1, (1, 0); Scenario 2, (0, 0); and Scenario 3, (1, 1).

b. Because pre-existing informative censoring is not present in Scenarios 1 and 2, it does not need to be considered.

c. Because Experiment F does not consider pre-existing censoring, no modelling is required.

d. Indicator terms for each day of the grace period; the model is fitted to records within the grace period, the only period during which artificial censoring can occur in the control arm.

e. In the intervention arm, artificial censoring can occur only on the final day of the grace period, and the model is fitted to records from that single day (Experiments A–D and F); time terms are therefore unnecessary, as the intercept absorbs the day-specific baseline hazard.

f. The separate model for pre-existing censoring includes no time terms in either arm; this is consistent with the data-generating mechanism, in which the pre-existing censoring hazard is constant over time given covariates.

g. In Experiment E, a single model combining artificial and pre-existing censoring is fitted to all person-time records over the entire follow-up. Both the control and intervention arms include no time terms.

IPCW, inverse probability of censoring weights
