## supplemental file 1 for "Pitfalls and Solutions in Clone-Censor-Weight for Target Trial Emulation: Insights from Review, Simulation, and Real-World Analyses"

**Supplementary file 1**

- Pages 3-5: Review of studies applying clone-censor-weight analysis to real-world medical data
- Pages 6-8: Details of the general principles of clone-censor-weight analysis
- Page 9: Figure S1. Directed acyclic graph used for simulation studies
- Page 10: Details of data generation in Simulation Study Part 1
- Pages 11-14: An intuitive explanation of the tail-heaviness index for inverse probability of censoring weights
- Pages 15-17: Details of clone-censor-weight analysis using real-world medical data
- Pages 18-19: Table S1. Patient Characteristics in Scenario 1 (A1D1), According to the Cutoff
- Pages 20-21: Table S2. Patient Characteristics in Scenario 2 (A1D1), According to the Cutoff
- Pages 22-23: Table S3. Patient Characteristics in Scenario 3 (A1D1), According to the Cutoff
- Pages 24-25: Table S4. The observed 30-day outcome incidences and the corresponding estimates from the ground-truth simulations and clone-censor-weight analyses in simulation Part 1
- Pages 26-27: Table S5. Patient Characteristics in Scenario 1 (A0.5D1), According to the Cutoff
- Pages 28-29: Table S6. Patient Characteristics in Scenario 1 (A1D0.5), According to the Cutoff
- Pages 30-31: Table S7. Patient Characteristics in Scenario 1 (A1D2), According to the Cutoff
- Pages 32-33: Table S8. Patient Characteristics in Scenario 1 (A1D4), According to the Cutoff
- Pages 34-35: Table S9. Patient Characteristics in Scenario 1 (A1D8), According to the Cutoff
- Pages 36-37: Table S10. Patient Characteristics in Scenario 1 (A2D1), According to the Cutoff
- Pages 38-39: Table S11. Patient Characteristics in Scenario 1 (A4D1), According to the Cutoff
- Pages 40-41: Table S12. Patient Characteristics in Scenario 1 (A8D1), According to the Cutoff
- Pages 42-43: Table S13. The observed 30-day outcome incidences and the corresponding estimates from the ground-truth simulations and clone-censor-weight analyses in simulation Part 2
- Page 44: Figure S2. Discrepancy between the oracle and estimated IPCW tail-heaviness indices
- Page 45: Results of simulations with varying magnitudes of covariate effects on the outcome
- Pages 46-48: Figure S3 Coverage probability according to the oracle IPCW tail-heaviness index, sample size, and standard deviation of the estimator under varying strengths of confounding affecting treatment assignment
- Pages 49-50: Detailed results of the clone-censor-weight analysis using real-world medical data
- Page 51: Figure S4. Patient flow
- Page 52: Table S14. Patient characteristics
- Page 53: Figure S5. IPCW-weighted Kaplan–Meier survival curves
- Page 54: Figure S6. Age distribution according to incident pre-existing censor status
- Pages 55-56: Explanation for counterintuitive results in Simulation Part 1
- Page 57: Discussion of the real-world medical data analysis results
- Page 58: References

**Review of studies applying clone-censor-weight analysis to real-world medical data**

To characterize how clone-censor-weight (CCW) analysis has been applied in practice, we searched PubMed in April 2026 using the following query: ("clone-censor-weighting"[All Fields] OR "cloning, censoring, and weighting"[All Fields] OR "clone-censor-weight"[All Fields] OR ("target trial emulation"[Title/Abstract] AND "clon*"[Title/Abstract] AND "censor*"[Title/Abstract])). The search identified 88 articles, of which 77 applied CCW analysis to real-world medical data. For each article, we extracted the title, PMID, publication year, and information on the following three aspects of CCW analysis.

First, we assessed whether time-varying covariates were included in the inverse probability of censoring weighting (IPCW) model. Specifically, we recorded whether the IPCW model used to address artificial censoring due to deviation from the assigned strategy incorporated covariates updated during follow-up rather than baseline covariates alone. A study was classified as “Yes” only when the IPCW model explicitly incorporated covariates updated during follow-up, as “No” when all covariates were measured at baseline, and as “Uncertain” when this could not be determined from the report.

Second, we assessed the handling of pre-existing censoring. We recorded whether the analysis accounted for the potential informativeness of censoring mechanisms other than artificial censoring, such as loss to follow-up, emigration, disenrollment, or transfer of care. Studies were classified as “Yes (IPCW)” when such censoring was addressed using IPCW, either in a model separate from or combined with the model for artificial censoring; “Yes (non-IPCW)” when it was addressed using another method; “No” when potentially informative pre-existing censoring occurred but its informativeness was not addressed; “Uncertain” when the handling could not be determined from the report; and “N/A” when no informative pre-existing censoring required adjustment. The “N/A” category included studies in which such censoring could not occur, follow-up was complete with no observed censoring, or the only pre-existing censoring was administrative censoring at the end of the study or at a fixed maximum follow-up time, which we regarded as non-informative.

Third, we assessed the handling of competing events. We recorded whether the analysis accounted for competing events relevant to the outcome under study. Studies were classified as “Yes (IPCW)” when competing events were addressed using IPCW; “Yes (non-IPCW)” when they were addressed using a competing-risk method, such as a Fine-Gray subdistribution hazard model, a cause-specific hazard model, or an Aalen-Johansen or weighted cumulative incidence estimator; “No” when a competing event could occur but was handled only as censoring; “Uncertain” when the handling could not be determined; and “N/A” when no competing event existed, such as when the outcome was all-cause mortality or a composite outcome including death, or when no competing event occurred in the cohort.

Because death may be treated either as a censoring event or as a competing event, we applied a consistent rule to avoid double-counting. When death was the outcome, including all-cause mortality, overall survival, or a component of a composite outcome, or when it was a competing event for a non-fatal outcome, its handling was assessed under the competing-event item, whereas the pre-existing-censoring item was assessed on the basis of non-death censoring only. An exception was made for studies that explicitly incorporated death as natural censoring within an IPCW model; in such cases, the handling of death was credited under the pre-existing-censoring item. When death was the only competing event and was fully accounted for in this manner, the competing-event item was classified as “N/A.”

Among the 77 studies that applied CCW analysis to real-world medical data, 34 (44.2%) incorporated time-varying covariates into the IPCW model, whereas 39 (50.6%) used baseline covariates only and 4 (5.2%) were uncertain. Among the 47 studies in which potentially informative pre-existing censoring required consideration, 7 (14.9%) addressed it using IPCW, 1 (2.1%) using a non-IPCW method, 37 (78.7%) did not address its potential informativeness, and 2 (4.3%) were uncertain. Among the 43 studies in which a competing event required consideration, 3 (7.0%) addressed it using IPCW, 16 (37.2%) using a non-IPCW competing-risk method, 22 (51.2%) did not account for the competing event, and 2 (4.7%) were uncertain.

The complete list of included articles is provided in Supplementary file 3.

**Details of the General Principles of Clone-Censor-Weight Analysis**

***Key Considerations in Specifying Treatment Strategies***

When conducting a CCW analysis, investigators must first specify the target trial protocol, including the treatment strategies to be compared. Treatments can generally be classified into two categories: point treatments, such as surgery, and sustained treatments, such as long-term medication. Treatment strategies are therefore commonly defined as follows:

1. **Initiation of treatment (point or sustained treatment):** When the clinical question is whether initiating a point or sustained treatment within the grace period is associated with better outcomes than not initiating it, a typical intervention strategy is “initiate treatment within the grace period,” whereas a typical control strategy is “do not initiate treatment within the grace period.”
2. **Initiation and continuation of sustained treatment:** When the clinical question is whether initiating a sustained treatment within the grace period and subsequently continuing it is associated with better outcomes than not initiating it, a typical intervention strategy is “initiate sustained treatment within the grace period and continue it thereafter,” whereas a typical control strategy is “do not initiate treatment within the grace period.”

Two important points should be considered when specifying treatment strategies. First, to uniquely define the potential outcomes—and therefore the causal effect—of interest, each treatment strategy must be specified unambiguously. For example, the intervention strategy in the first case can be defined more precisely as follows: “Initiate treatment on the final day of the grace period if the patient has not already initiated treatment.” Such clarification is important for accurately interpreting the estimated effect. Second, treatment strategies should be carefully defined with consideration of other factors or interventions that may affect both treatment decisions and outcomes, which may require more detailed specifications. For example, suppose an investigator is interested in whether bronchial artery embolization (BAE), a catheter-based treatment for severe hemoptysis, performed within 5 days of hospitalization improves in-hospital survival among patients with severe hemoptysis. Lung-resection surgery is another treatment option for severe hemoptysis, although it is performed less frequently than BAE. If the investigator wishes to estimate the effect of BAE without incorporating the effect of surgery, the intervention strategy may be defined as “initiate BAE within 5 days without undergoing surgery,” and the control strategy as “initiate neither BAE nor surgery within 5 days.” Procedures performed after day 5 would not be restricted by these strategy definitions, although alternative strategies could incorporate such procedures if necessary. In this study, we focused on the first type of strategy, treatment initiation, because it represents the most fundamental application of CCW analysis. In addition, this strategy has been adopted in most published studies using CCW analysis.

***Description of Each Component of Clone-Censor-Weight Analysis***

Step 1. Clone

At time zero, each eligible patient is replicated once for each predefined treatment strategy. For example, if two strategies are prespecified, each patient is duplicated into two clones, with one clone assigned to each strategy.

Step 2. Censor

Each clone is artificially censored at the time it deviates from its assigned treatment strategy. For example:

- A patient undergoes BAE on day 3 without undergoing surgery. The clone assigned to the intervention strategy (“BAE within 5 days without surgery”) remains uncensored because the patient adheres to this strategy. In contrast, the clone assigned to the control strategy (“no BAE or surgery within 5 days”) is artificially censored on day 3.
- Another patient undergoes neither BAE nor surgery within 5 days. The clone assigned to the intervention strategy is artificially censored at day 5, whereas the clone assigned to the control strategy remains uncensored.

After the clone and censor steps, the resulting dataset represents follow-up under the predefined treatment strategies, although artificial censoring may introduce selection bias that must subsequently be addressed through weighting. Standard causal-effect estimation methods can then be applied to the appropriately weighted data.

From a technical perspective, investigators should organize the data in a time-split format, such as by day, week, or month. The dataset should include indicator variables such as outcome, pre_censor, and artificial_censor, each coded as 1 at the time when the outcome, pre-existing censoring, or artificial censoring occurs, respectively. If competing events are present and are accounted for in the analysis, an additional variable, such as competing_outcome, should also be created.

Step 3. Weight

IPCW is used to account for the probability of remaining uncensored. In practice, the procedure is as follows:

1. An IPCW model is fitted with artificial censoring as the dependent variable and covariates that may affect censoring as predictors.
2. For each individual at each time point, the cumulative probability of remaining uncensored through that time is estimated using the IPCW model, and the inverse of this probability is calculated.
3. The resulting weights are incorporated into the analysis, typically through weighted Kaplan-Meier estimation or weighted outcome models.

Because per-protocol effects represent the outcomes that would be expected if individuals adhered to their assigned treatment strategies from time zero onward, this weighting step is essential. Without appropriate adjustment, artificial censoring due to deviation from the assigned strategy can introduce selection bias. For example, if older patients are more likely to undergo BAE early, the mean age among patients remaining uncensored under the control strategy (“no BAE or surgery within 5 days”) may decrease from 65 years at baseline to 62, 60, 57, and 52 years on subsequent days. IPCW addresses this selection bias by assigning greater weights to uncensored individuals who have a higher predicted probability of being censored—in this example, older patients who remain uncensored.

**Figure S1. Directed acyclic graph used for simulation studies**

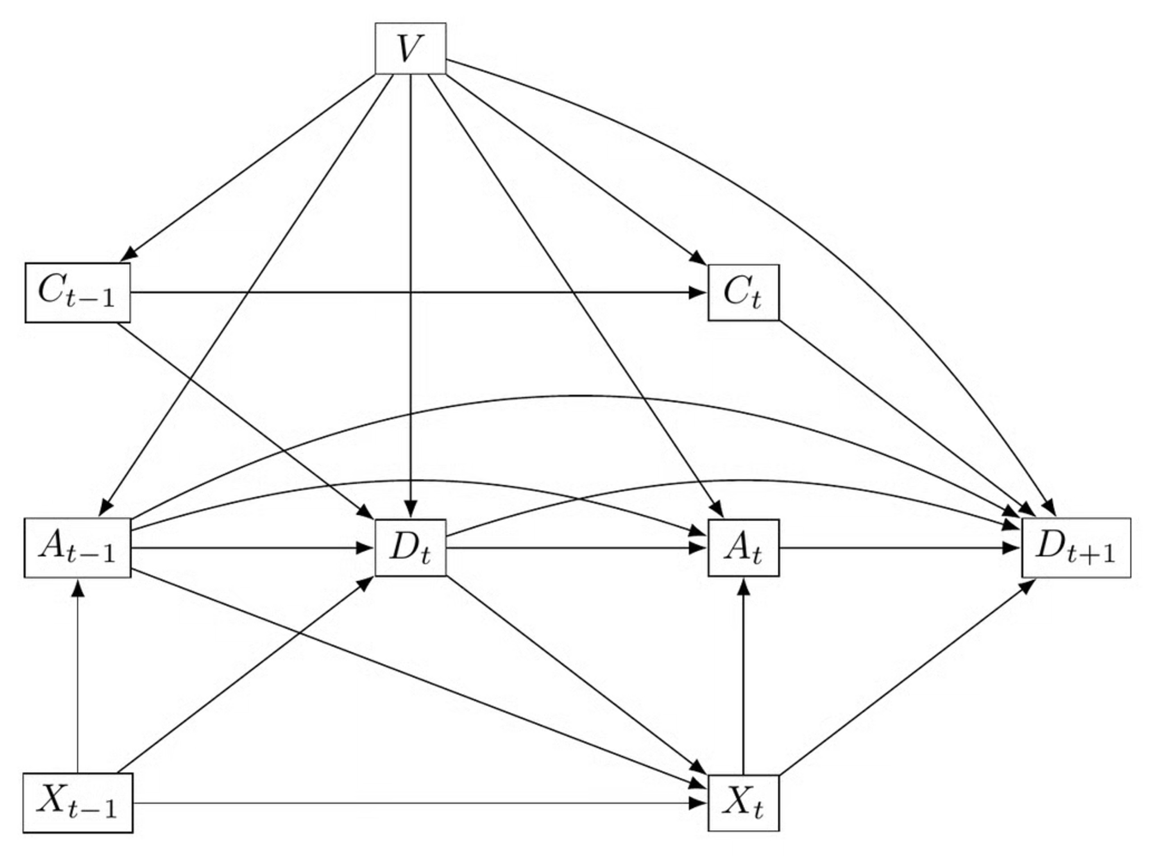

This directed acyclic graph corresponds to Scenario 3, which includes three baseline covariates (age, sex, and Charlson Comorbidity Index), one time-varying covariate (SpO2, blood oxygen saturation), treatment, outcome (in-hospital death), and pre-existing censoring exist. $V$, $X_{t}$, $A_{t}$, $D_{t}$, $C_{t}$ denote baseline covariates, time-varying covariate, treatment, outcome, and pre-existing censoring, respectively. Variable with the subscript “$t$” represent their status at time $t$. In Scenario 1, pre-existing censoring $C$ is absent, whereas in Scenario 2, the arrow from time-varying covariate $X$ to treatment $A$ is also absent.

**Details of data generation in Simulation Study Part 1**

The simulated datasets included six variables: three baseline covariates (age, sex, and CCI), one time-varying covariate (SpO₂), treatment, and the outcome of in-hospital death. In Scenario 3, which involved pre-existing censoring, an additional variable indicating pre-existing censoring was generated. Age was modeled as a continuous integer variable, sex as a binary variable (male or female), and CCI as a categorical variable with values of 0, 1, or 2. CCI is a comorbidity burden index, with higher values indicating a greater burden of comorbidities. Although the CCI can take values greater than 2, we restricted its range to 0–2 for simplicity. SpO₂ was modeled as a continuous variable ranging from 0 to 1, representing blood oxygen saturation, and is generally associated with the prognosis of respiratory disease.

The treatment of interest was more likely to be administered to older patients, female patients, patients with higher CCI categories, and patients with lower SpO₂ values. The outcome was more likely to occur in older patients, male patients, patients with higher CCI categories, and patients with lower SpO₂ values. Pre-existing censoring, which may be interpreted as discharge alive, was more likely to occur in younger patients, female patients, and patients with lower CCI categories. SpO₂ affected the outcome, but it was assumed not to affect pre-existing censoring. The magnitudes of the effects of age, sex, and CCI on the outcome and pre-existing censoring were specified separately. The daily probability of treatment initiation varied with time since baseline (declining exponentially over follow-up) as well as with covariates, whereas the hazard of pre-existing censoring was constant over time given covariates.

In the ground-truth datasets, all individuals were forced to adhere to each predefined treatment strategy. For example, under the intervention strategy of “initiating treatment within the grace period,” any individual who had not initiated treatment by the day before the final day of the grace period was assigned treatment with probability 1 on the final day.

Further details are provided in Supplementary file 2.

**An intuitive explanation of the tail-heaviness index for inverse probability of censoring weights**

***Why extreme weights are a concern***

IPCW requires each individual who remains uncensored to “stand in” for similar individuals who were censored. To do this, each uncensored individual is assigned a weight equal to the inverse of their estimated probability of remaining uncensored. A well-known concern with this approach is the occurrence of extremely large weights. For example, if one patient receives a weight larger than the combined weight of all other patients, the estimate may be driven largely by that single patient.

However, the more fundamental problem is not simply whether an extreme weight happens to appear in a particular dataset, but whether the underlying weight distribution has a heavy tail^1^. As the sample size increases, the empirical distribution of the observed weights more closely approximates the underlying distribution from which the data are sampled. For example, if 10% of individuals in the underlying distribution have weights of 3.0 or greater, the observed proportion may vary substantially in a small sample but will tend to approach 10% as the sample size increases. When the underlying distribution has a light tail, a sufficiently large sample therefore reflects a distribution in which individuals with large weights are sufficiently uncommon that they do not dominate the analysis. Consequently, the estimator generally converges in the usual manner as the sample size increases. In contrast, when the underlying distribution has a heavy tail, a large sample likewise more closely reflects the underlying distribution, but this distribution itself assigns sufficient probability to large weights that individuals with such weights can continue to exert a dominant influence on the analysis even in large samples. Consequently, the estimator may fail to stabilize at the usual rate, and confidence intervals may have substantially lower coverage than their nominal level.

Because tail heaviness is a property of the underlying distribution, it cannot be reliably assessed simply by inspecting the largest observed weights in one dataset. We therefore need a diagnostic that directly describes how heavy the tail is. The tail-heaviness index is intended to serve this purpose.

***Heavy tails and the tail index α***

The “tail” of a distribution describes how frequently extremely large values occur. Human height, for example, has a light tail: values several times the average height essentially do not occur. Income has a much heavier tail: most incomes are within an ordinary range, but values tens, hundreds, or even thousands of times larger can occur.

A standard mathematical model for a heavy tail is the Pareto-type, or power-law, tail, in which the probability of observing a value larger than $z$ approximately follows

$$P\left( Z>z \right)\approx C\cdot z^{-\alpha}.$$

The exponent $\alpha$, called the tail index, provides a simple measure of tail heaviness: smaller values of $\alpha$ indicate a heavier tail and therefore a greater tendency to produce extreme values.

Two thresholds are particularly useful for understanding this idea. When $\alpha$ $\leq2$, the variance of a Pareto-type distribution is infinite, meaning that averages do not stabilize at the usual rate as the sample size increases. When $\alpha$ $\leq1$, even the mean is infinite, and extremely large observations can dominate the distribution regardless of how large the sample becomes. Intuitively, $\alpha$ describes a balance between how rare extreme observations are and how large they can become: the smaller $\alpha$ is, the more likely it is that a few exceptionally large observations will dominate.

On a log–log plot, a Pareto-type tail approximately forms a straight line with slope $-\alpha$. The Hill estimator is a standard method for estimating $\alpha$ from the largest observations in the data.

***What the tail-heaviness index computes***

Our diagnostic applies this idea to the components that build up the IPCW weights over time, rather than only examining the final weights.

During follow-up, an individual's IPCW is updated at each time point. The existing weight is multiplied by a new factor: the inverse of the estimated conditional probability of remaining uncensored at that time point, given that the individual has remained uncensored up to the previous time point.

At each time point within the grace period, we examine these new inverse-probability factors among individuals who are still at risk. A large new factor is particularly influential when it occurs in an individual who has already accumulated a large weight from earlier time points. We therefore give greater importance to such individuals by weighting them according to the square of their accumulated inverse uncensoring probability up to the previous time point. The square is used because variance calculations depend on squared weights.

We then apply a weighted Hill estimator to the largest current inverse-probability factors. This gives an estimated tail index $\alpha$ for each treatment strategy and each time point. Because instability at even a single time point can affect the overall analysis, we use the minimum index across time points—the point with the heaviest estimated tail—as a summary measure for each strategy.

Although $\alpha=2$ is the familiar threshold at which the variance of an ordinary Pareto-type variable becomes infinite, our tail-heaviness index is instead interpreted using a threshold of 1. The reason is that our diagnostic is relevant to the variance of the IPCW estimator. Consequently, the question becomes whether the mean of this variance-related quantity is finite, rather than whether its variance is finite. For a Pareto-type tail, this mean is finite when $\alpha>1$.

Thus, an index above 1 is consistent with a finite variance-related moment, whereas an index below 1 warns that this moment may be infinite and conventional confidence intervals may become unreliable. However, because $\alpha$ is itself estimated with uncertainty and the Pareto model is only an approximation to the true tail, an estimated value below 1 should be interpreted as a warning sign rather than definitive proof of infinite variance.

We also separately examine whether the weight accumulated from previous time points is itself excessively heavy-tailed. This diagnostic is not reported in the main manuscript because, in our simulations, it did not identify any additional problematic settings: whenever its estimated index was below 1, the reported current-component index was also below1

Further details, including the mathematical formulation, are provided in Supplementary file 2.

**Details of clone-censor-weight analysis using real-world medical data**

***Data Source***

We used the Japanese Diagnosis Procedure Combination (DPC) database, a nationwide inpatient database covering more than 1000 acute-care hospitals, representing approximately 90% of all tertiary care hospitals in Japan^2^. The database includes detailed information on patient demographics, diagnoses coded based on International Classification of Diseases, 10th Revision, (ICD-10) codes, medical procedures (including surgeries), daily drug administration, and discharge status. A validation study of this database reported diagnostic sensitivity and specificity of 78.9% and 93.2%, respectively, with >90% accuracy for medical procedures^3^.

***Eligibility criteria***

We included patients admitted with hemoptysis between July 1, 2010, and March 31, 2023, who met the following criteria: 1) confirmed diagnosis of hemoptysis (ICD-10 codes R042, R048, and R049) as the main diagnosis or admission–precipitating diagnosis, 2) age ≥18 years, and 3) ventilation use on the day of admission. For patients meeting these criteria multiple times, only the first episode was analyzed.

The DPC database records clinical events on a daily basis and does not provide hourly information. Therefore, the exact time of death could not be determined for patients who died on a given day. When a patient received any intervention on the recorded date of death—such as oral medication, intravenous infusion, testing, a medical procedure, surgery, or a meal—indicating that the patient had survived for at least part of that day, death was treated as having occurred on the following day for the purposes of this analysis. This approach allowed for the possibility that interventions could exert clinical effects within several hours. No corresponding adjustment was made for variables other than death, for which the original recorded dates were used.

***Treatment strategies***

We defined the following two treatment strategies: 1) intervention strategy: BAE within 5 days of admission without lung-resection surgery; and 2) control strategy: No BAE or lung-resection surgery within 5 days. We excluded lung-resection surgery in these strategies because both BAE and lung-resection surgery are major treatment options for severe hemoptysis, and inclusion of surgery would complicate the estimation.

***Assignment procedure***

We used clone-censor-weight (CCW) analysis. Each eligible patient was cloned once for each predefined treatment strategy, and one clone was assigned to each strategy.

***Outcomes***

The outcome of interest was 30-day all-cause in-hospital mortality.

***Follow-up period***

Follow-up began on the date of admission (day 0) and continued until the earliest occurrence of death, discharge alive from the hospital, or day 30. Discharge alive was treated as pre-existing censoring. We considered this censoring potentially informative because treatment for hemoptysis was likely to have been completed by the time of discharge and patients in better clinical condition were more likely to be discharged.

***Causal contrast***

CCW analysis estimates a per-protocol effect representing the contrast between outcomes under full adherence to the predefined treatment strategies. We therefore estimated the per-protocol effect of each strategy.

***Analysis plan***

We conducted the CCW analysis in three steps. In the clone step, each patient was cloned across the two predefined treatment strategies. In the censor step, each clone was artificially censored when the patient deviated from the assigned strategy. In the weight step, two IPCW models were fitted separately for each strategy: one for artificial censoring and another for pre-existing censoring. Both models included all covariates described below. For each clone at each time point, the final weight was calculated as the product of the weights obtained from the two IPCW models.

We used weighted Kaplan-Meier estimators to estimate the 30-day risk of all-cause in-hospital mortality under each strategy. We performed 1000 bootstrap iterations to obtain 95% confidence intervals (CIs) for the estimated outcome risk under each strategy. Risk differences (RDs) and risk ratios (RRs), with their corresponding 95% CIs, were also estimated to compare the two strategies.

Baseline time-fixed covariates included age, sex, and bronchopulmonary carcinoma as the cause of hemoptysis, which has been associated with particularly poor prognosis^4^. Time-varying covariates included intravenous tranexamic acid use, blood transfusion, bronchoscopy, intensive care unit admission, mechanical ventilation, and contrast-enhanced computed tomography. Each time-varying covariate was coded as 0 (absent) or 1 (present) and updated daily. Because this analysis was intended primarily to illustrate the application of CCW analysis, we included only covariates considered particularly important.

To assess the potential validity of statistical inference, we calculated the IPCW tail-heaviness index for each strategy, as described in the main manuscript. We also estimated bootstrap standard errors for mortality under each strategy and for the RD and RR using the 1000 bootstrap iterations.

***Exploratory analysis***

In the primary analysis, we accounted for potentially informative pre-existing censoring. In the exploratory analysis, we deliberately omitted this adjustment to demonstrate the effect of not accounting for pre-existing censoring. Specifically, we constructed an IPCW model that accounted only for artificial censoring.

To characterize differences between patients who remained hospitalized and those who were discharged alive and therefore experienced pre-existing censoring, we also compared the age distributions of these two groups.

**Table S1. Patient Characteristics in Scenario 1 (A1D1), According to the Cutoff**

| **Cutoff 0** |  |  |  |
| --- | --- | --- | --- |
| **Characteristic** | **Overall** | **Intervention** | **Control** |
| Age, median [IQR] | 74.1 [58.6, 83.4] | 79.7 [70.8, 87.1] | 73.5 [57.5, 83.0] |
| Male sex, % | 50.0% | 52.8% | 49.8% |
| CCI, median [IQR] | 1.0 [0.1, 2.0] | 1.8 [1.0, 2.0] | 1.0 [0.0, 2.0] |
| Baseline SpO₂, median [IQR] | 0.890 [0.864, 0.914] | 0.872 [0.845, 0.896] | 0.892 [0.866, 0.915] |
| SpO₂ at the cutoff, median [IQR] | 0.890 [0.864, 0.914] | 0.872 [0.845, 0.896] | 0.892 [0.866, 0.915] |
| **Cutoff 2** |  |  |  |
| **Characteristic** | **Overall** | **Intervention** | **Control** |
| Age, median [IQR] | 74.1 [58.6, 83.4] | 78.5 [69.0, 86.1] | 72.9 [56.3, 82.7] |
| Male sex, % | 50.0% | 50.1% | 50.0% |
| CCI, median [IQR] | 1.0 [0.1, 2.0] | 1.1 [1.0, 2.0] | 1.0 [0.0, 2.0] |
| Baseline SpO₂, median [IQR] | 0.890 [0.864, 0.914] | 0.877 [0.851, 0.900] | 0.893 [0.867, 0.916] |
| SpO₂ at the cutoff, median [IQR] | 0.911 [0.873, 0.941] | 0.901 [0.869, 0.927] | 0.915 [0.875, 0.943] |
| **Cutoff 4** |  |  |  |
| **Characteristic** | **Overall** | **Intervention** | **Control** |
| Age, median [IQR] | 74.1 [58.6, 83.4] | 77.7 [67.6, 85.5] | 72.8 [55.9, 82.6] |
| Male sex, % | 50.0% | 48.7% | 50.4% |
| CCI, median [IQR] | 1.0 [0.1, 2.0] | 1.0 [1.0, 2.0] | 1.0 [0.0, 2.0] |
| Baseline SpO₂, median [IQR] | 0.890 [0.864, 0.914] | 0.880 [0.855, 0.902] | 0.894 [0.867, 0.917] |
| SpO₂ at the cutoff, median [IQR] | 0.921 [0.882, 0.950] | 0.906 [0.875, 0.933] | 0.928 [0.886, 0.954] |

Intervention and control refer to patients who received the intervention within the grace period, including the cutoff day, and those who did not, respectively. A1D1 represents the reference setting for the magnitude of confounding. In AxDy, x and y indicate the factors by which the effects of the confounders on intervention assignment (A) and the outcome (D), respectively, were multiplied relative to the reference setting. For simplicity, patient characteristics are presented only for a sample size of 10 000. Age, CCI, baseline SpO₂, and SpO₂ at the cutoff are summarized as medians [IQRs], whereas male sex is presented as a percentage. All values represent averages across the 1000 simulation iterations.

CCI, Charlson Comorbidity Index; IQR, interquartile range; SpO₂, peripheral oxygen saturation.

**Table S2. Patient Characteristics in Scenario 2 (A1D1), According to the Cutoff**

| **Cutoff 0** |  |  |  |
| --- | --- | --- | --- |
| **Characteristic** | **Overall** | **Intervention** | **Control** |
| Age, median [IQR] | 74.1 [58.6, 83.4] | 78.5 [68.3, 86.4] | 73.7 [57.9, 83.1] |
| Male sex, % | 50.0% | 50.6% | 50.0% |
| CCI, median [IQR] | 1.0 [0.1, 2.0] | 1.5 [1.0, 2.0] | 1.0 [0.0, 2.0] |
| Baseline SpO₂, median [IQR] | 0.890 [0.864, 0.914] | 0.886 [0.860, 0.909] | 0.891 [0.864, 0.914] |
| SpO₂ at the cutoff, median [IQR] | 0.890 [0.864, 0.914] | 0.886 [0.860, 0.909] | 0.891 [0.864, 0.914] |
| **Cutoff 2** |  |  |  |
| **Characteristic** | **Overall** | **Intervention** | **Control** |
| Age, median [IQR] | 74.1 [58.6, 83.4] | 77.5 [66.6, 85.6] | 73.3 [57.1, 82.9] |
| Male sex, % | 50.0% | 48.5% | 50.3% |
| CCI, median [IQR] | 1.0 [0.1, 2.0] | 1.0 [1.0, 2.0] | 1.0 [0.0, 2.0] |
| Baseline SpO₂, median [IQR] | 0.890 [0.864, 0.914] | 0.890 [0.865, 0.912] | 0.891 [0.864, 0.914] |
| SpO₂ at the cutoff, median [IQR] | 0.912 [0.873, 0.941] | 0.917 [0.888, 0.941] | 0.911 [0.869, 0.941] |
| **Cutoff 4** |  |  |  |
| **Characteristic** | **Overall** | **Intervention** | **Control** |
| Age, median [IQR] | 74.1 [58.6, 83.4] | 76.8 [65.4, 85.0] | 73.2 [56.7, 82.8] |
| Male sex, % | 50.0% | 47.3% | 50.7% |
| CCI, median [IQR] | 1.0 [0.1, 2.0] | 1.0 [1.0, 2.0] | 1.0 [0.0, 2.0] |
| Baseline SpO₂, median [IQR] | 0.890 [0.864, 0.914] | 0.892 [0.867, 0.913] | 0.890 [0.863, 0.914] |
| SpO₂ at the cutoff, median [IQR] | 0.923 [0.883, 0.951] | 0.924 [0.894, 0.949] | 0.922 [0.878, 0.951] |

Intervention and control refer to patients who received the intervention within the grace period, including the cutoff day, and those who did not, respectively. A1D1 represents the reference setting for the magnitude of confounding. In AxDy, x and y indicate the factors by which the effects of the confounders on intervention assignment (A) and the outcome (D), respectively, were multiplied relative to the reference setting. For simplicity, patient characteristics are presented only for a sample size of 10 000. Age, CCI, baseline SpO₂, and SpO₂ at the cutoff are summarized as medians [IQRs], whereas male sex is presented as a percentage. All values represent averages across the 1000 simulation iterations.

CCI, Charlson Comorbidity Index; IQR, interquartile range; SpO₂, peripheral oxygen saturation.

**Table S3. Patient Characteristics in Scenario 3 (A1D1), According to the Cutoff**

| **Cutoff 0** |  |  |  |
| --- | --- | --- | --- |
| **Characteristic** | **Overall** | **Intervention** | **Control** |
| Age, median [IQR] | 74.1 [58.6, 83.4] | 79.7 [70.8, 87.1] | 73.5 [57.5, 83.0] |
| Male sex, % | 50.0% | 52.9% | 49.8% |
| CCI, median [IQR] | 1.0 [0.1, 2.0] | 1.8 [1.0, 2.0] | 1.0 [0.0, 2.0] |
| Baseline SpO₂, median [IQR] | 0.890 [0.864, 0.914] | 0.872 [0.845, 0.896] | 0.892 [0.866, 0.915] |
| SpO₂ at the cutoff, median [IQR] | 0.890 [0.864, 0.914] | 0.872 [0.845, 0.896] | 0.892 [0.866, 0.915] |
| **Cutoff 2** |  |  |  |
| **Characteristic** | **Overall** | **Intervention** | **Control** |
| Age, median [IQR] | 74.1 [58.6, 83.4] | 78.5 [69.0, 86.1] | 72.9 [56.3, 82.7] |
| Male sex, % | 50.0% | 50.2% | 50.0% |
| CCI, median [IQR] | 1.0 [0.1, 2.0] | 1.1 [1.0, 2.0] | 1.0 [0.0, 2.0] |
| Baseline SpO₂, median [IQR] | 0.890 [0.864, 0.914] | 0.877 [0.851, 0.900] | 0.893 [0.867, 0.916] |
| SpO₂ at the cutoff, median [IQR] | 0.911 [0.873, 0.941] | 0.901 [0.869, 0.927] | 0.915 [0.875, 0.943] |
| **Cutoff 4** |  |  |  |
| **Characteristic** | **Overall** | **Intervention** | **Control** |
| Age, median [IQR] | 74.1 [58.6, 83.4] | 77.7 [67.6, 85.5] | 72.8 [55.9, 82.6] |
| Male sex, % | 50.0% | 48.7% | 50.4% |
| CCI, median [IQR] | 1.0 [0.1, 2.0] | 1.0 [1.0, 2.0] | 1.0 [0.0, 2.0] |
| Baseline SpO₂, median [IQR] | 0.890 [0.864, 0.914] | 0.880 [0.855, 0.902] | 0.893 [0.867, 0.917] |
| SpO₂ at the cutoff, median [IQR] | 0.921 [0.882, 0.950] | 0.906 [0.875, 0.933] | 0.928 [0.886, 0.954] |

Intervention and control refer to patients who received the intervention within the grace period, including the cutoff day, and those who did not, respectively. A1D1 represents the reference setting for the magnitude of confounding. In AxDy, x and y indicate the factors by which the effects of the confounders on intervention assignment (A) and the outcome (D), respectively, were multiplied relative to the reference setting. For simplicity, patient characteristics are presented only for a sample size of 10 000. Age, CCI, baseline SpO₂, and SpO₂ at the cutoff are summarized as medians [IQRs], whereas male sex is presented as a percentage. All values represent averages across the 1000 simulation iterations.

CCI, Charlson Comorbidity Index; IQR, interquartile range; SpO₂, peripheral oxygen saturation.

**Table S4. The observed 30-day outcome incidences and the corresponding estimates from the ground-truth simulations and clone-censor-weight analyses in simulation Part 1**

|  | Observed (raw) | Observed (raw) | Ground truth simulation | Ground truth simulation | CCW simulation | CCW simulation |
| --- | --- | --- | --- | --- | --- | --- |
| **Arm** | Intervention^a^ | control^a^ | Intervention^b^ | control^b^ | Intervention^c^ | control^c^ |
| **Scenario 1 (a1d1) experimentA cut0** | 0.264 (0.016) | 0.369 (0.005) | 0.155 [0.154, 0.155] | 0.382 [0.381, 0.382] | 0.155 (0.010) | 0.381 (0.005) |
| **Scenario 1 (a1d1) experimentB cut0** | 0.264 (0.016) | 0.369 (0.005) | 0.155 [0.154, 0.155] | 0.382 [0.381, 0.382] | 0.155 (0.010) | 0.381 (0.005) |
| **Scenario 1 (a1d1) experimentA cut2** | 0.238 (0.010) | 0.385 (0.005) | 0.222 [0.222, 0.223] | 0.413 [0.413, 0.413] | 0.223 (0.008) | 0.413 (0.005) |
| **Scenario 1 (a1d1) experimentB cut2** | 0.238 (0.010) | 0.385 (0.005) | 0.222 [0.222, 0.223] | 0.413 [0.413, 0.413] | 0.247 (0.010) | 0.402 (0.005) |
| **Scenario 1 (a1d1) experimentA cut4** | 0.213 (0.008) | 0.402 (0.006) | 0.262 [0.262, 0.262] | 0.435 [0.434, 0.435] | 0.262 (0.008) | 0.434 (0.005) |
| **Scenario 1 (a1d1) experimentB cut4** | 0.213 (0.008) | 0.402 (0.006) | 0.262 [0.262, 0.262] | 0.435 [0.434, 0.435] | 0.280 (0.010) | 0.412 (0.006) |
| **Scenario 2 (a1d1) experimentC cut0** | 0.197 (0.015) | 0.399 (0.005) | 0.155 [0.154, 0.155] | 0.403 [0.403, 0.403] | 0.154 (0.011) | 0.403 (0.005) |
| **Scenario 2 (a1d1) experimentC cut2** | 0.162 (0.009) | 0.426 (0.005) | 0.226 [0.226, 0.226] | 0.428 [0.427, 0.428] | 0.226 (0.010) | 0.428 (0.005) |
| **Scenario 2 (a1d1) experimentC cut4** | 0.139 (0.008) | 0.450 (0.006) | 0.271 [0.270, 0.271] | 0.444 [0.444, 0.444] | 0.270 (0.009) | 0.444 (0.006) |
| **Scenario 3 (a1d1) experimentD cut0** | 0.264 (0.016) | 0.369 (0.005) | 0.155 [0.155, 0.155] | 0.382 [0.381, 0.382] | 0.156 (0.012) | 0.382 (0.006) |
| **Scenario 3 (a1d1) experimentE cut0** | 0.264 (0.016) | 0.369 (0.005) | 0.155 [0.155, 0.155] | 0.382 [0.381, 0.382] | 0.224 (0.303) | 0.382 (0.006) |
| **Scenario 3 (a1d1) experimentF cut0** | 0.264 (0.016) | 0.369 (0.005) | 0.155 [0.155, 0.155] | 0.382 [0.381, 0.382] | 0.181 (0.012) | 0.407 (0.006) |
| **Scenario 3 (a1d1) experimentD cut2** | 0.238 (0.010) | 0.386 (0.005) | 0.222 [0.222, 0.222] | 0.413 [0.412, 0.413] | 0.225 (0.012) | 0.413 (0.006) |
| **Scenario 3 (a1d1) experimentE cut2** | 0.238 (0.010) | 0.386 (0.005) | 0.222 [0.222, 0.222] | 0.413 [0.412, 0.413] | 0.220 (0.013) | 0.416 (0.008) |
| **Scenario 3 (a1d1) experimentF cut2** | 0.238 (0.010) | 0.386 (0.005) | 0.222 [0.222, 0.222] | 0.413 [0.412, 0.413] | 0.243 (0.011) | 0.439 (0.006) |
| **Scenario 3 (a1d1) experimentD cut4** | 0.212 (0.009) | 0.402 (0.005) | 0.262 [0.262, 0.262] | 0.435 [0.434, 0.435] | 0.264 (0.010) | 0.435 (0.007) |
| **Scenario 3 (a1d1) experimentE cut4** | 0.212 (0.009) | 0.402 (0.005) | 0.262 [0.262, 0.262] | 0.435 [0.434, 0.435] | 0.264 (0.007) | 0.441 (0.010) |
| **Scenario 3 (a1d1) experimentF cut4** | 0.212 (0.009) | 0.402 (0.005) | 0.262 [0.262, 0.262] | 0.435 [0.434, 0.435] | 0.282 (0.010) | 0.463 (0.006) |

a. For the observed (raw) data, the intervention and control groups comprise simulated patients who received the intervention within the grace period, including the cutoff day, and those who did not, respectively. Values are presented as the mean (standard deviation) across 1000 simulation replicates.

b. The ground-truth outcome incidences for the intervention and control strategies were obtained by independently generating 10 000 000 individuals from the structural causal model and deterministically assigning the treatment variable according to each strategy. The ground-truth value was calculated as the proportion of simulated individuals who experienced the outcome by day 30. Values are presented with Wald 95% confidence intervals.

c. For the CCW analyses, the intervention and control estimates were derived from clones assigned to the corresponding treatment strategies. Values are presented as the mean (standard deviation) across 1000 simulation replicates.

A1D1 represents the reference setting for the magnitude of confounding. In AxDy, x and y indicate the factors by which the effects of the confounders on intervention assignment (A) and the outcome (D), respectively, were multiplied relative to the reference setting. For simplicity, results are presented only for a sample size of 10 000.

CCW, clone-censor-weight.

**Table S5. Patient Characteristics in Scenario 1 (A0.5D1), According to the Cutoff**

| **Cutoff 0** |  |  |  |
| --- | --- | --- | --- |
| **Characteristic** | **Overall** | **Intervention** | **Control** |
| Age, median [IQR] | 74.1 [58.6, 83.4] | 77.5 [66.5, 85.5] | 73.8 [58.1, 83.2] |
| Male sex, % | 50.0% | 51.4% | 49.9% |
| CCI, median [IQR] | 1.0 [0.1, 2.0] | 1.0 [1.0, 2.0] | 1.0 [0.0, 2.0] |
| Baseline SpO₂, median [IQR] | 0.890 [0.864, 0.914] | 0.880 [0.853, 0.904] | 0.891 [0.865, 0.914] |
| SpO₂ at the cutoff, median [IQR] | 0.890 [0.864, 0.914] | 0.880 [0.853, 0.904] | 0.891 [0.865, 0.914] |
| **Cutoff 2** |  |  |  |
| **Characteristic** | **Overall** | **Intervention** | **Control** |
| Age, median [IQR] | 74.1 [58.6, 83.4] | 76.3 [64.6, 84.6] | 73.7 [57.7, 83.1] |
| Male sex, % | 50.0% | 49.2% | 50.1% |
| CCI, median [IQR] | 1.0 [0.1, 2.0] | 1.0 [1.0, 2.0] | 1.0 [0.0, 2.0] |
| Baseline SpO₂, median [IQR] | 0.890 [0.864, 0.914] | 0.884 [0.859, 0.907] | 0.891 [0.865, 0.915] |
| SpO₂ at the cutoff, median [IQR] | 0.912 [0.872, 0.941] | 0.909 [0.878, 0.934] | 0.912 [0.871, 0.942] |
| **Cutoff 4** |  |  |  |
| **Characteristic** | **Overall** | **Intervention** | **Control** |
| Age, median [IQR] | 74.1 [58.6, 83.4] | 75.5 [63.0, 84.0] | 73.7 [57.6, 83.2] |
| Male sex, % | 50.0% | 48.0% | 50.5% |
| CCI, median [IQR] | 1.0 [0.1, 2.0] | 1.0 [1.0, 2.0] | 1.0 [0.0, 2.0] |
| Baseline SpO₂, median [IQR] | 0.890 [0.864, 0.914] | 0.887 [0.862, 0.909] | 0.891 [0.864, 0.915] |
| SpO₂ at the cutoff, median [IQR] | 0.922 [0.882, 0.950] | 0.915 [0.884, 0.941] | 0.925 [0.882, 0.953] |

Intervention and control refer to patients who received the intervention within the grace period, including the cutoff day, and those who did not, respectively. A1D1 represents the reference setting for the magnitude of confounding. In AxDy, x and y indicate the factors by which the effects of the confounders on intervention assignment (A) and the outcome (D), respectively, were multiplied relative to the reference setting. For simplicity, patient characteristics are presented only for a sample size of 10 000. Age, CCI, baseline SpO₂, and SpO₂ at the cutoff are summarized as medians [IQRs], whereas male sex is presented as a percentage. All values represent averages across the 1000 simulation iterations.

CCI, Charlson Comorbidity Index; IQR, interquartile range; SpO₂, peripheral oxygen saturation.

**Table S6. Patient Characteristics in Scenario 1 (A1D0.5), According to the Cutoff**

| **Cutoff 0** |  |  |  |
| --- | --- | --- | --- |
| **Characteristic** | **Overall** | **Intervention** | **Control** |
| Age, median [IQR] | 74.1 [58.6, 83.4] | 79.7 [70.8, 87.1] | 73.5 [57.5, 83.0] |
| Male sex, % | 50.0% | 52.8% | 49.8% |
| CCI, median [IQR] | 1.0 [0.1, 2.0] | 1.8 [1.0, 2.0] | 1.0 [0.0, 2.0] |
| Baseline SpO₂, median [IQR] | 0.890 [0.864, 0.914] | 0.872 [0.845, 0.896] | 0.892 [0.866, 0.915] |
| SpO₂ at the cutoff, median [IQR] | 0.890 [0.864, 0.914] | 0.872 [0.845, 0.896] | 0.892 [0.866, 0.915] |
| **Cutoff 2** |  |  |  |
| **Characteristic** | **Overall** | **Intervention** | **Control** |
| Age, median [IQR] | 74.1 [58.6, 83.4] | 79.2 [70.1, 86.7] | 72.6 [55.8, 82.4] |
| Male sex, % | 50.0% | 52.2% | 49.5% |
| CCI, median [IQR] | 1.0 [0.1, 2.0] | 1.5 [1.0, 2.0] | 1.0 [0.0, 2.0] |
| Baseline SpO₂, median [IQR] | 0.890 [0.864, 0.914] | 0.874 [0.847, 0.898] | 0.894 [0.868, 0.917] |
| SpO₂ at the cutoff, median [IQR] | 0.906 [0.865, 0.938] | 0.896 [0.861, 0.924] | 0.910 [0.866, 0.941] |
| **Cutoff 4** |  |  |  |
| **Characteristic** | **Overall** | **Intervention** | **Control** |
| Age, median [IQR] | 74.1 [58.6, 83.4] | 78.8 [69.4, 86.4] | 71.9 [54.7, 82.0] |
| Male sex, % | 50.0% | 51.7% | 49.4% |
| CCI, median [IQR] | 1.0 [0.1, 2.0] | 1.2 [1.0, 2.0] | 1.0 [0.0, 2.0] |
| Baseline SpO₂, median [IQR] | 0.890 [0.864, 0.914] | 0.875 [0.849, 0.899] | 0.895 [0.870, 0.918] |
| SpO₂ at the cutoff, median [IQR] | 0.913 [0.869, 0.946] | 0.898 [0.864, 0.928] | 0.920 [0.871, 0.951] |

Intervention and control refer to patients who received the intervention within the grace period, including the cutoff day, and those who did not, respectively. A1D1 represents the reference setting for the magnitude of confounding. In AxDy, x and y indicate the factors by which the effects of the confounders on intervention assignment (A) and the outcome (D), respectively, were multiplied relative to the reference setting. For simplicity, patient characteristics are presented only for a sample size of 10 000. Age, CCI, baseline SpO₂, and SpO₂ at the cutoff are summarized as medians [IQRs], whereas male sex is presented as a percentage. All values represent averages across the 1000 simulation iterations.

CCI, Charlson Comorbidity Index; IQR, interquartile range; SpO₂, peripheral oxygen saturation.

**Table S7. Patient Characteristics in Scenario 1 (A1D2), According to the Cutoff**

| **Cutoff 0** |  |  |  |
| --- | --- | --- | --- |
| **Characteristic** | **Overall** | **Intervention** | **Control** |
| Age, median [IQR] | 74.1 [58.6, 83.4] | 79.7 [70.8, 87.1] | 73.5 [57.5, 83.0] |
| Male sex, % | 50.0% | 52.8% | 49.8% |
| CCI, median [IQR] | 1.0 [0.1, 2.0] | 1.8 [1.0, 2.0] | 1.0 [0.0, 2.0] |
| Baseline SpO₂, median [IQR] | 0.890 [0.864, 0.914] | 0.872 [0.845, 0.896] | 0.892 [0.866, 0.915] |
| SpO₂ at the cutoff, median [IQR] | 0.890 [0.864, 0.914] | 0.872 [0.845, 0.896] | 0.892 [0.866, 0.915] |
| **Cutoff 2** |  |  |  |
| **Characteristic** | **Overall** | **Intervention** | **Control** |
| Age, median [IQR] | 74.1 [58.6, 83.4] | 76.9 [66.1, 85.1] | 73.6 [57.4, 83.1] |
| Male sex, % | 50.0% | 47.0% | 50.4% |
| CCI, median [IQR] | 1.0 [0.1, 2.0] | 1.0 [1.0, 2.0] | 1.0 [0.0, 2.0] |
| Baseline SpO₂, median [IQR] | 0.890 [0.864, 0.914] | 0.883 [0.857, 0.905] | 0.892 [0.865, 0.915] |
| SpO₂ at the cutoff, median [IQR] | 0.924 [0.892, 0.948] | 0.916 [0.891, 0.937] | 0.926 [0.893, 0.950] |
| **Cutoff 4** |  |  |  |
| **Characteristic** | **Overall** | **Intervention** | **Control** |
| Age, median [IQR] | 74.1 [58.6, 83.4] | 75.6 [63.5, 84.1] | 73.8 [57.7, 83.2] |
| Male sex, % | 50.0% | 45.1% | 50.9% |
| CCI, median [IQR] | 1.0 [0.1, 2.0] | 1.0 [1.0, 2.0] | 1.0 [0.0, 2.0] |
| Baseline SpO₂, median [IQR] | 0.890 [0.864, 0.914] | 0.887 [0.861, 0.909] | 0.891 [0.864, 0.915] |
| SpO₂ at the cutoff, median [IQR] | 0.935 [0.903, 0.957] | 0.922 [0.897, 0.945] | 0.938 [0.906, 0.960] |

Intervention and control refer to patients who received the intervention within the grace period, including the cutoff day, and those who did not, respectively. A1D1 represents the reference setting for the magnitude of confounding. In AxDy, x and y indicate the factors by which the effects of the confounders on intervention assignment (A) and the outcome (D), respectively, were multiplied relative to the reference setting. For simplicity, patient characteristics are presented only for a sample size of 10 000. Age, CCI, baseline SpO₂, and SpO₂ at the cutoff are summarized as medians [IQRs], whereas male sex is presented as a percentage. All values represent averages across the 1000 simulation iterations.

CCI, Charlson Comorbidity Index; IQR, interquartile range; SpO₂, peripheral oxygen saturation.

**Table S8. Patient Characteristics in Scenario 1 (A1D4), According to the Cutoff**

| **Cutoff 0** |  |  |  |
| --- | --- | --- | --- |
| **Characteristic** | **Overall** | **Intervention** | **Control** |
| Age, median [IQR] | 74.1 [58.6, 83.4] | 79.7 [70.8, 87.1] | 73.5 [57.5, 83.0] |
| Male sex, % | 50.0% | 52.8% | 49.8% |
| CCI, median [IQR] | 1.0 [0.1, 2.0] | 1.8 [1.0, 2.0] | 1.0 [0.0, 2.0] |
| Baseline SpO₂, median [IQR] | 0.890 [0.864, 0.914] | 0.872 [0.845, 0.896] | 0.892 [0.866, 0.915] |
| SpO₂ at the cutoff, median [IQR] | 0.890 [0.864, 0.914] | 0.872 [0.845, 0.896] | 0.892 [0.866, 0.915] |
| **Cutoff 2** |  |  |  |
| **Characteristic** | **Overall** | **Intervention** | **Control** |
| Age, median [IQR] | 74.1 [58.6, 83.4] | 76.8 [64.8, 85.2] | 73.7 [57.9, 83.1] |
| Male sex, % | 50.0% | 47.7% | 50.3% |
| CCI, median [IQR] | 1.0 [0.1, 2.0] | 1.0 [1.0, 2.0] | 1.0 [0.0, 2.0] |
| Baseline SpO₂, median [IQR] | 0.890 [0.864, 0.914] | 0.883 [0.854, 0.907] | 0.891 [0.865, 0.914] |
| SpO₂ at the cutoff, median [IQR] | 0.933 [0.904, 0.954] | 0.928 [0.906, 0.946] | 0.933 [0.904, 0.955] |
| **Cutoff 4** |  |  |  |
| **Characteristic** | **Overall** | **Intervention** | **Control** |
| Age, median [IQR] | 74.1 [58.6, 83.4] | 75.2 [61.2, 84.3] | 73.9 [58.2, 83.2] |
| Male sex, % | 50.0% | 46.0% | 50.5% |
| CCI, median [IQR] | 1.0 [0.1, 2.0] | 1.0 [0.9, 2.0] | 1.0 [0.0, 2.0] |
| Baseline SpO₂, median [IQR] | 0.890 [0.864, 0.914] | 0.887 [0.858, 0.911] | 0.891 [0.865, 0.914] |
| SpO₂ at the cutoff, median [IQR] | 0.942 [0.914, 0.962] | 0.932 [0.910, 0.952] | 0.944 [0.915, 0.963] |

Intervention and control refer to patients who received the intervention within the grace period, including the cutoff day, and those who did not, respectively. A1D1 represents the reference setting for the magnitude of confounding. In AxDy, x and y indicate the factors by which the effects of the confounders on intervention assignment (A) and the outcome (D), respectively, were multiplied relative to the reference setting. For simplicity, patient characteristics are presented only for a sample size of 10 000. Age, CCI, baseline SpO₂, and SpO₂ at the cutoff are summarized as medians [IQRs], whereas male sex is presented as a percentage. All values represent averages across the 1000 simulation iterations.

CCI, Charlson Comorbidity Index; IQR, interquartile range; SpO₂, peripheral oxygen saturation.

**Table S9. Patient Characteristics in Scenario 1 (A1D8), According to the Cutoff**

| **Cutoff 0** |  |  |  |
| --- | --- | --- | --- |
| **Characteristic** | **Overall** | **Intervention** | **Control** |
| Age, median [IQR] | 74.1 [58.6, 83.4] | 79.7 [70.8, 87.1] | 73.5 [57.5, 83.0] |
| Male sex, % | 50.0% | 52.8% | 49.8% |
| CCI, median [IQR] | 1.0 [0.1, 2.0] | 1.8 [1.0, 2.0] | 1.0 [0.0, 2.0] |
| Baseline SpO₂, median [IQR] | 0.890 [0.864, 0.914] | 0.872 [0.845, 0.896] | 0.892 [0.866, 0.915] |
| SpO₂ at the cutoff, median [IQR] | 0.890 [0.864, 0.914] | 0.872 [0.845, 0.896] | 0.892 [0.866, 0.915] |
| **Cutoff 2** |  |  |  |
| **Characteristic** | **Overall** | **Intervention** | **Control** |
| Age, median [IQR] | 74.1 [58.6, 83.4] | 77.2 [65.0, 85.6] | 73.7 [57.9, 83.1] |
| Male sex, % | 50.0% | 48.9% | 50.1% |
| CCI, median [IQR] | 1.0 [0.1, 2.0] | 1.0 [1.0, 2.0] | 1.0 [0.0, 2.0] |
| Baseline SpO₂, median [IQR] | 0.890 [0.864, 0.914] | 0.880 [0.851, 0.906] | 0.891 [0.865, 0.914] |
| SpO₂ at the cutoff, median [IQR] | 0.936 [0.909, 0.956] | 0.933 [0.912, 0.950] | 0.937 [0.909, 0.957] |
| **Cutoff 4** |  |  |  |
| **Characteristic** | **Overall** | **Intervention** | **Control** |
| Age, median [IQR] | 74.1 [58.6, 83.4] | 75.8 [61.3, 84.8] | 73.9 [58.3, 83.2] |
| Male sex, % | 50.0% | 47.4% | 50.3% |
| CCI, median [IQR] | 1.0 [0.1, 2.0] | 1.0 [1.0, 2.0] | 1.0 [0.0, 2.0] |
| Baseline SpO₂, median [IQR] | 0.890 [0.864, 0.914] | 0.884 [0.855, 0.910] | 0.891 [0.865, 0.914] |
| SpO₂ at the cutoff, median [IQR] | 0.945 [0.918, 0.964] | 0.937 [0.916, 0.956] | 0.946 [0.919, 0.965] |

Intervention and control refer to patients who received the intervention within the grace period, including the cutoff day, and those who did not, respectively. A1D1 represents the reference setting for the magnitude of confounding. In AxDy, x and y indicate the factors by which the effects of the confounders on intervention assignment (A) and the outcome (D), respectively, were multiplied relative to the reference setting. For simplicity, patient characteristics are presented only for a sample size of 10 000. Age, CCI, baseline SpO₂, and SpO₂ at the cutoff are summarized as medians [IQRs], whereas male sex is presented as a percentage. All values represent averages across the 1000 simulation iterations.

CCI, Charlson Comorbidity Index; IQR, interquartile range; SpO₂, peripheral oxygen saturation.

**Table S10. Patient Characteristics in Scenario 1 (A2D1), According to the Cutoff**

| **Cutoff 0** |  |  |  |
| --- | --- | --- | --- |
| **Characteristic** | **Overall** | **Intervention** | **Control** |
| Age, median [IQR] | 74.1 [58.6, 83.4] | 81.8 [74.6, 88.7] | 72.2 [55.5, 82.0] |
| Male sex, % | 50.0% | 54.8% | 49.2% |
| CCI, median [IQR] | 1.0 [0.1, 2.0] | 2.0 [1.0, 2.0] | 1.0 [0.0, 2.0] |
| Baseline SpO₂, median [IQR] | 0.890 [0.864, 0.914] | 0.861 [0.835, 0.885] | 0.895 [0.870, 0.917] |
| SpO₂ at the cutoff, median [IQR] | 0.890 [0.864, 0.914] | 0.861 [0.835, 0.885] | 0.895 [0.870, 0.917] |
| **Cutoff 2** |  |  |  |
| **Characteristic** | **Overall** | **Intervention** | **Control** |
| Age, median [IQR] | 74.1 [58.6, 83.4] | 80.5 [72.7, 87.5] | 70.4 [52.8, 81.0] |
| Male sex, % | 50.0% | 51.8% | 49.3% |
| CCI, median [IQR] | 1.0 [0.1, 2.0] | 2.0 [1.0, 2.0] | 1.0 [0.0, 2.0] |
| Baseline SpO₂, median [IQR] | 0.890 [0.864, 0.914] | 0.869 [0.844, 0.891] | 0.898 [0.874, 0.920] |
| SpO₂ at the cutoff, median [IQR] | 0.911 [0.876, 0.940] | 0.892 [0.862, 0.918] | 0.920 [0.884, 0.946] |
| **Cutoff 4** |  |  |  |
| **Characteristic** | **Overall** | **Intervention** | **Control** |
| Age, median [IQR] | 74.1 [58.6, 83.4] | 79.7 [71.4, 86.9] | 69.8 [51.9, 80.7] |
| Male sex, % | 50.0% | 50.3% | 49.9% |
| CCI, median [IQR] | 1.0 [0.1, 2.0] | 1.9 [1.0, 2.0] | 1.0 [0.0, 2.0] |
| Baseline SpO₂, median [IQR] | 0.890 [0.864, 0.914] | 0.873 [0.847, 0.895] | 0.899 [0.875, 0.921] |
| SpO₂ at the cutoff, median [IQR] | 0.919 [0.882, 0.948] | 0.897 [0.868, 0.924] | 0.933 [0.897, 0.957] |

Intervention and control refer to patients who received the intervention within the grace period, including the cutoff day, and those who did not, respectively. A1D1 represents the reference setting for the magnitude of confounding. In AxDy, x and y indicate the factors by which the effects of the confounders on intervention assignment (A) and the outcome (D), respectively, were multiplied relative to the reference setting. For simplicity, patient characteristics are presented only for a sample size of 10 000. Age, CCI, baseline SpO₂, and SpO₂ at the cutoff are summarized as medians [IQRs], whereas male sex is presented as a percentage. All values represent averages across the 1000 simulation iterations.

CCI, Charlson Comorbidity Index; IQR, interquartile range; SpO₂, peripheral oxygen saturation.

**Table S11. Patient Characteristics in Scenario 1 (A4D1), According to the Cutoff**

| **Cutoff 0** |  |  |  |
| --- | --- | --- | --- |
| **Characteristic** | **Overall** | **Intervention** | **Control** |
| Age, median [IQR] | 74.1 [58.6, 83.4] | 82.1 [75.5, 88.7] | 67.6 [50.5, 78.7] |
| Male sex, % | 50.0% | 55.2% | 47.5% |
| CCI, median [IQR] | 1.0 [0.1, 2.0] | 2.0 [1.0, 2.0] | 1.0 [0.0, 2.0] |
| Baseline SpO₂, median [IQR] | 0.890 [0.864, 0.914] | 0.861 [0.837, 0.882] | 0.903 [0.882, 0.922] |
| SpO₂ at the cutoff, median [IQR] | 0.890 [0.864, 0.914] | 0.861 [0.837, 0.882] | 0.903 [0.882, 0.922] |
| **Cutoff 2** |  |  |  |
| **Characteristic** | **Overall** | **Intervention** | **Control** |
| Age, median [IQR] | 74.1 [58.6, 83.4] | 80.6 [73.3, 87.5] | 63.3 [46.9, 76.1] |
| Male sex, % | 50.0% | 52.9% | 47.4% |
| CCI, median [IQR] | 1.0 [0.1, 2.0] | 2.0 [1.0, 2.0] | 1.0 [0.0, 2.0] |
| Baseline SpO₂, median [IQR] | 0.890 [0.864, 0.914] | 0.869 [0.845, 0.890] | 0.908 [0.888, 0.926] |
| SpO₂ at the cutoff, median [IQR] | 0.911 [0.881, 0.939] | 0.895 [0.869, 0.918] | 0.929 [0.900, 0.951] |
| **Cutoff 4** |  |  |  |
| **Characteristic** | **Overall** | **Intervention** | **Control** |
| Age, median [IQR] | 74.1 [58.6, 83.4] | 80.0 [72.3, 87.0] | 62.2 [46.0, 75.6] |
| Male sex, % | 50.0% | 52.1% | 47.7% |
| CCI, median [IQR] | 1.0 [0.1, 2.0] | 2.0 [1.0, 2.0] | 1.0 [0.0, 1.8] |
| Baseline SpO₂, median [IQR] | 0.890 [0.864, 0.914] | 0.872 [0.847, 0.893] | 0.909 [0.889, 0.927] |
| SpO₂ at the cutoff, median [IQR] | 0.917 [0.884, 0.946] | 0.899 [0.871, 0.924] | 0.940 [0.909, 0.960] |

Intervention and control refer to patients who received the intervention within the grace period, including the cutoff day, and those who did not, respectively. A1D1 represents the reference setting for the magnitude of confounding. In AxDy, x and y indicate the factors by which the effects of the confounders on intervention assignment (A) and the outcome (D), respectively, were multiplied relative to the reference setting. For simplicity, patient characteristics are presented only for a sample size of 10 000. Age, CCI, baseline SpO₂, and SpO₂ at the cutoff are summarized as medians [IQRs], whereas male sex is presented as a percentage. All values represent averages across the 1000 simulation iterations.

CCI, Charlson Comorbidity Index; IQR, interquartile range; SpO₂, peripheral oxygen saturation.

**Table S12. Patient Characteristics in Scenario 1 (A8D1), According to the Cutoff**

| **Cutoff 0** |  |  |  |
| --- | --- | --- | --- |
| **Characteristic** | **Overall** | **Intervention** | **Control** |
| Age, median [IQR] | 74.1 [58.6, 83.4] | 81.0 [74.2, 87.7] | 59.9 [45.2, 73.1] |
| Male sex, % | 50.0% | 53.9% | 45.9% |
| CCI, median [IQR] | 1.0 [0.1, 2.0] | 2.0 [1.0, 2.0] | 1.0 [0.0, 1.0] |
| Baseline SpO₂, median [IQR] | 0.890 [0.864, 0.914] | 0.868 [0.845, 0.888] | 0.913 [0.895, 0.929] |
| SpO₂ at the cutoff, median [IQR] | 0.890 [0.864, 0.914] | 0.868 [0.845, 0.888] | 0.913 [0.895, 0.929] |
| **Cutoff 2** |  |  |  |
| **Characteristic** | **Overall** | **Intervention** | **Control** |
| Age, median [IQR] | 74.1 [58.6, 83.4] | 79.8 [72.2, 86.8] | 55.6 [42.6, 70.0] |
| Male sex, % | 50.0% | 52.4% | 46.1% |
| CCI, median [IQR] | 1.0 [0.1, 2.0] | 2.0 [1.0, 2.0] | 1.0 [0.0, 1.0] |
| Baseline SpO₂, median [IQR] | 0.890 [0.864, 0.914] | 0.874 [0.850, 0.894] | 0.916 [0.899, 0.932] |
| SpO₂ at the cutoff, median [IQR] | 0.913 [0.886, 0.939] | 0.902 [0.877, 0.924] | 0.935 [0.909, 0.955] |
| **Cutoff 4** |  |  |  |
| **Characteristic** | **Overall** | **Intervention** | **Control** |
| Age, median [IQR] | 74.1 [58.6, 83.4] | 79.4 [71.4, 86.5] | 54.9 [42.0, 69.6] |
| Male sex, % | 50.0% | 52.1% | 46.3% |
| CCI, median [IQR] | 1.0 [0.1, 2.0] | 1.5 [1.0, 2.0] | 1.0 [0.0, 1.0] |
| Baseline SpO₂, median [IQR] | 0.890 [0.864, 0.914] | 0.875 [0.851, 0.896] | 0.917 [0.900, 0.933] |
| SpO₂ at the cutoff, median [IQR] | 0.918 [0.887, 0.946] | 0.904 [0.877, 0.930] | 0.944 [0.916, 0.963] |

Intervention and control refer to patients who received the intervention within the grace period, including the cutoff day, and those who did not, respectively. A1D1 represents the reference setting for the magnitude of confounding. In AxDy, x and y indicate the factors by which the effects of the confounders on intervention assignment (A) and the outcome (D), respectively, were multiplied relative to the reference setting. For simplicity, patient characteristics are presented only for a sample size of 10 000. Age, CCI, baseline SpO₂, and SpO₂ at the cutoff are summarized as medians [IQRs], whereas male sex is presented as a percentage. All values represent averages across the 1000 simulation iterations.

CCI, Charlson Comorbidity Index; IQR, interquartile range; SpO₂, peripheral oxygen saturation.

**Table S13. The observed 30-day outcome incidences and the corresponding estimates from the ground-truth simulations and clone-censor-weight analyses in simulation Part 2**

|  | Observed (raw) | Observed (raw) | Ground truth simulation | Ground truth simulation | CCW simulation | CCW simulation |
| --- | --- | --- | --- | --- | --- | --- |
| **Arm** | Intervention^a^ | control^a^ | Intervention^b^ | control^b^ | Intervention^c^ | control^c^ |
| **Scenario 1 (a1d1) experimentA cut0** | 0.264 (0.016) | 0.369 (0.005) | 0.155 [0.154, 0.155] | 0.382 [0.381, 0.382] | 0.155 (0.010) | 0.381 (0.005) |
| **Scenario 1 (a1d1) experimentA cut2** | 0.238 (0.010) | 0.385 (0.005) | 0.222 [0.222, 0.223] | 0.413 [0.413, 0.413] | 0.223 (0.008) | 0.413 (0.005) |
| **Scenario 1 (a1d1) experimentA cut4** | 0.213 (0.008) | 0.402 (0.006) | 0.262 [0.262, 0.262] | 0.435 [0.434, 0.435] | 0.262 (0.008) | 0.434 (0.005) |
| **Scenario 1 (a0.5d1) experimentA cut0** | 0.211 (0.017) | 0.400 (0.005) | 0.155 [0.154, 0.155] | 0.406 [0.405, 0.406] | 0.155 (0.011) | 0.405 (0.005) |
| **Scenario 1 (a0.5d1) experimentA cut2** | 0.184 (0.011) | 0.421 (0.005) | 0.226 [0.226, 0.227] | 0.430 [0.429, 0.430] | 0.226 (0.010) | 0.430 (0.005) |
| **Scenario 1 (a0.5d1) experimentA cut4** | 0.162 (0.009) | 0.439 (0.005) | 0.271 [0.271, 0.272] | 0.446 [0.446, 0.446] | 0.271 (0.009) | 0.446 (0.005) |
| **Scenario 1 (a2d1) experimentA cut0** | 0.336 (0.013) | 0.289 (0.005) | 0.155 [0.154, 0.155] | 0.326 [0.326, 0.327] | 0.155 (0.010) | 0.326 (0.005) |
| **Scenario 1 (a2d1) experimentA cut2** | 0.302 (0.009) | 0.294 (0.005) | 0.208 [0.208, 0.209] | 0.373 [0.372, 0.373] | 0.210 (0.011) | 0.373 (0.006) |
| **Scenario 1 (a2d1) experimentA cut4** | 0.275 (0.008) | 0.306 (0.006) | 0.234 [0.234, 0.235] | 0.406 [0.405, 0.406] | 0.236 (0.012) | 0.406 (0.007) |
| **Scenario 1 (a4d1) experimentA cut0** | 0.334 (0.008) | 0.158 (0.005) | 0.155 [0.154, 0.155] | 0.259 [0.259, 0.260] | 0.158 (0.019) | 0.259 (0.010) |
| **Scenario 1 (a4d1) experimentA cut2** | 0.288 (0.007) | 0.149 (0.005) | 0.179 [0.179, 0.180] | 0.319 [0.319, 0.320] | 0.189 (0.026) | 0.312 (0.054) |
| **Scenario 1 (a4d1) experimentA cut4** | 0.270 (0.006) | 0.156 (0.005) | 0.188 [0.188, 0.188] | 0.364 [0.364, 0.364] | 0.200 (0.024) | 0.343 (0.071) |
| **Scenario 1 (a8d1) experimentA cut0** | 0.277 (0.006) | 0.074 (0.004) | 0.155 [0.154, 0.155] | 0.228 [0.228, 0.228] | 0.171 (0.030) | 0.209 (0.100) |
| **Scenario 1 (a8d1) experimentA cut2** | 0.245 (0.006) | 0.072 (0.004) | 0.162 [0.162, 0.162] | 0.294 [0.293, 0.294] | 0.191 (0.031) | 0.181 (0.121) |
| **Scenario 1 (a8d1) experimentA cut4** | 0.236 (0.005) | 0.075 (0.005) | 0.165 [0.165, 0.165] | 0.341 [0.340, 0.341] | 0.195 (0.025) | 0.194 (0.125) |
| **Scenario 1 (a1d0.5) experimentA cut0** | 0.080 (0.010) | 0.200 (0.004) | 0.052 [0.052, 0.052] | 0.206 [0.206, 0.206] | 0.052 (0.007) | 0.206 (0.004) |
| **Scenario 1 (a1d0.5) experimentA cut2** | 0.081 (0.006) | 0.216 (0.005) | 0.079 [0.079, 0.080] | 0.233 [0.233, 0.233] | 0.079 (0.007) | 0.233 (0.005) |
| **Scenario 1 (a1d0.5) experimentA cut4** | 0.078 (0.005) | 0.230 (0.005) | 0.100 [0.100, 0.101] | 0.257 [0.257, 0.257] | 0.101 (0.007) | 0.257 (0.005) |
| **Scenario 1 (a1d2) experimentA cut0** | 0.566 (0.017) | 0.550 (0.005) | 0.368 [0.367, 0.368] | 0.565 [0.565, 0.565] | 0.368 (0.013) | 0.565 (0.005) |
| **Scenario 1 (a1d2) experimentA cut2** | 0.449 (0.014) | 0.566 (0.005) | 0.458 [0.458, 0.459] | 0.580 [0.579, 0.580] | 0.458 (0.008) | 0.580 (0.005) |
| **Scenario 1 (a1d2) experimentA cut4** | 0.391 (0.013) | 0.579 (0.005) | 0.492 [0.492, 0.493] | 0.588 [0.588, 0.589] | 0.492 (0.007) | 0.588 (0.005) |
| **Scenario 1 (a1d4) experimentA cut0** | 0.772 (0.015) | 0.673 (0.005) | 0.562 [0.561, 0.562] | 0.687 [0.687, 0.688] | 0.561 (0.018) | 0.687 (0.005) |
| **Scenario 1 (a1d4) experimentA cut2** | 0.640 (0.015) | 0.685 (0.005) | 0.625 [0.624, 0.625] | 0.694 [0.694, 0.694] | 0.624 (0.007) | 0.694 (0.005) |
| **Scenario 1 (a1d4) experimentA cut4** | 0.576 (0.015) | 0.694 (0.005) | 0.647 [0.647, 0.648] | 0.698 [0.697, 0.698] | 0.647 (0.006) | 0.697 (0.005) |
| **Scenario 1 (a1d8) experimentA cut0** | 0.860 (0.013) | 0.740 (0.004) | 0.671 [0.671, 0.672] | 0.753 [0.752, 0.753] | 0.671 (0.019) | 0.752 (0.004) |
| **Scenario 1 (a1d8) experimentA cut2** | 0.750 (0.015) | 0.749 (0.004) | 0.709 [0.709, 0.709] | 0.756 [0.756, 0.757] | 0.708 (0.007) | 0.756 (0.004) |
| **Scenario 1 (a1d8) experimentA cut4** | 0.692 (0.014) | 0.755 (0.004) | 0.725 [0.725, 0.725] | 0.759 [0.758, 0.759] | 0.725 (0.006) | 0.758 (0.004) |

a. For the observed (raw) data, the intervention and control groups comprise simulated patients who received the intervention within the grace period, including the cutoff day, and those who did not, respectively. Values are presented as the mean (standard deviation) across 1000 simulation replicates.

b. The ground-truth outcome incidences for the intervention and control strategies were obtained by independently generating 10 000 000 individuals from the structural causal model and deterministically assigning the treatment variable according to each strategy. The ground-truth value was calculated as the proportion of simulated individuals who experienced the outcome by day 30. Values are presented with Wald 95% confidence intervals.

c. For the CCW analyses, the intervention and control estimates were derived from clones assigned to the corresponding treatment strategies. Values are presented as the mean (standard deviation) across 1000 simulation replicates.

A1D1 represents the reference setting for the magnitude of confounding. In AxDy, x and y indicate the factors by which the effects of the confounders on intervention assignment (A) and the outcome (D), respectively, were multiplied relative to the reference setting. For simplicity, results are presented only for a sample size of 10 000.

CCW, clone-censor-weight.

**Figure S2. Discrepancy between the oracle and estimated IPCW tail-heaviness indices**

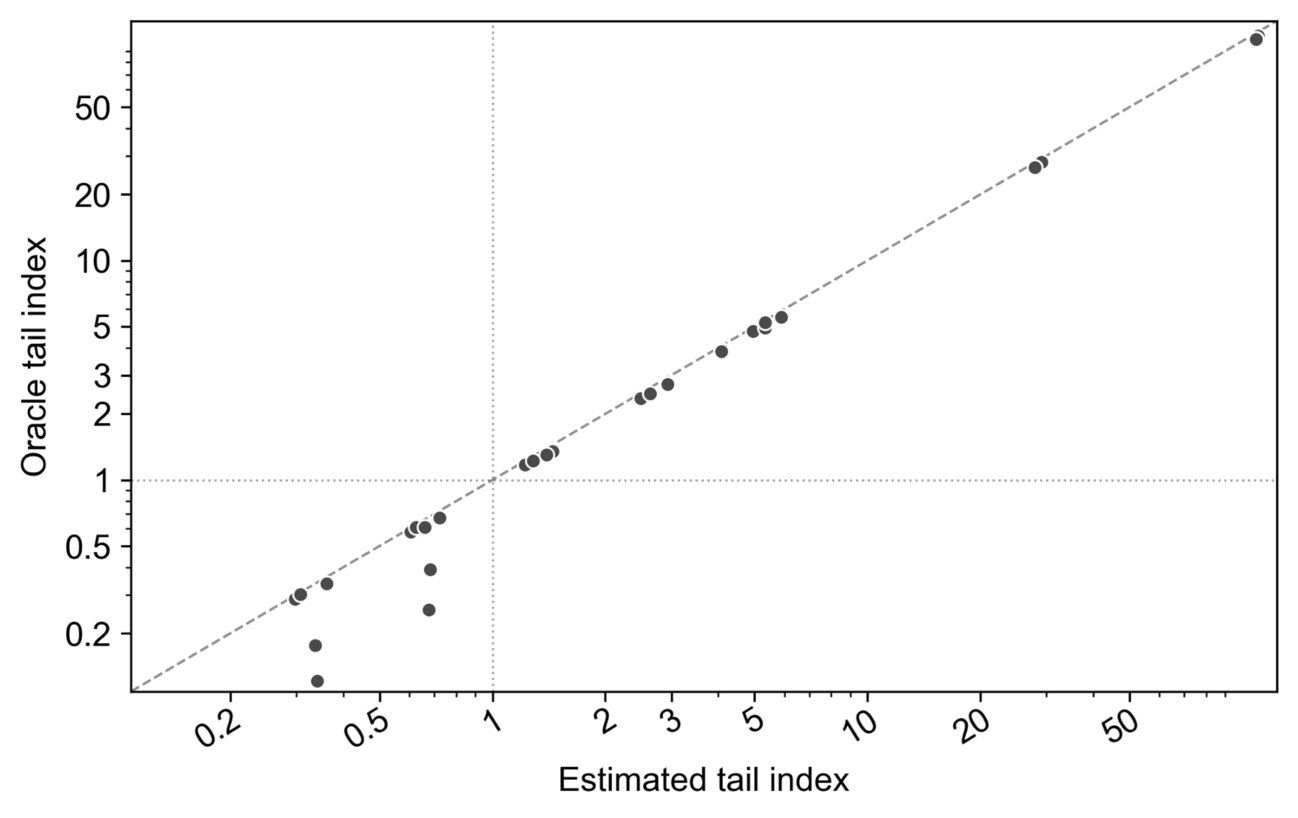

This figure shows the discrepancy between the oracle IPCW tail-heaviness index and the corresponding index estimated from the fitted IPCW models.

IPCW, inverse probability of censoring weighting.

**Results of simulations with varying magnitudes of covariate effects on the outcome**

Fig. S3 presents the results obtained when the strength of the associations between the covariates and the outcome was varied. Fig. S3 panels (a) and (b) show the relationships between the oracle tail-heaviness index and coverage probability for the control and intervention strategies, respectively, at a sample size of 10 000. For both strategies, all settings had an index of ≥1 and achieved coverage close to the nominal level of 0.95. Fig. 3 panel (c) shows how coverage varied according to sample size and the oracle tail-heaviness index. Across all settings, coverage increased with sample size and approached 0.95 at a sample size of 10 000. Fig. 3 panel (d) shows the relationship between the estimator's standard deviation and coverage, stratified by the oracle tail-heaviness index. All experiments had an index of ≥1, and coverage increased as the estimator's standard deviation decreased. Specifically, the nominal coverage of 0.95 was not achieved in any experiment with a standard deviation of ≥0.10, whereas it was approached in experiments with a standard deviation below 0.01.

**Figure S3. Coverage probability according to the oracle IPCW tail-heaviness index, sample size, and standard deviation of the estimator under varying strengths of confounding affecting outcome occurrence**

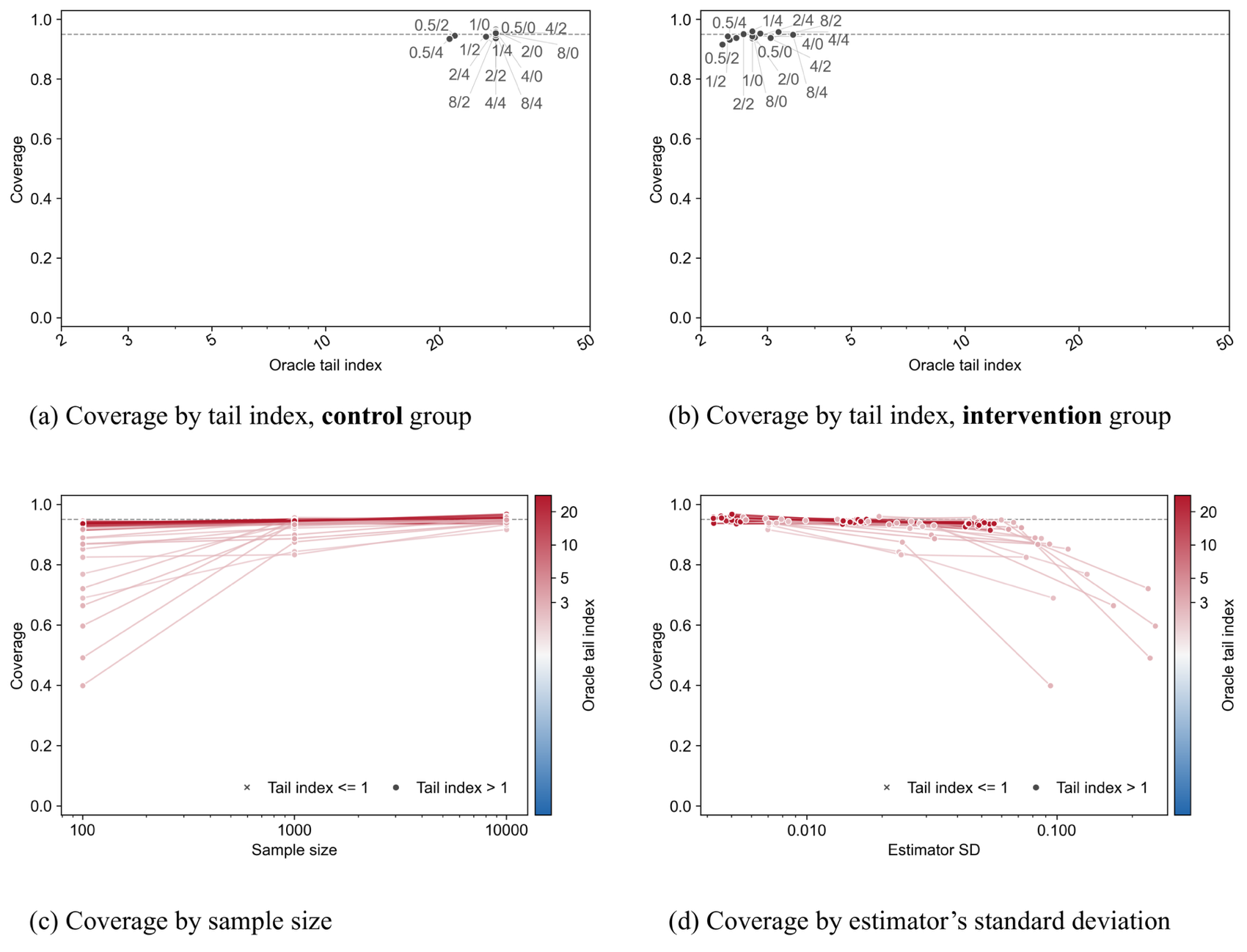

This figure shows coverage probability when the magnitude of the covariate effects on treatment assignment was varied. Panels (a) and (b) show the relationship between the oracle IPCW tail-heaviness index and coverage probability under the control and intervention strategies, respectively. In labels of the form “x/y” (e.g., 4/2), x denotes the factor by which the covariate effects on outcome occurrence were multiplied relative to the reference setting used in Part 1, and y denotes the cutoff day, defined as the final day of the grace period. Panel (c) shows coverage probability across sample sizes, stratified by the oracle IPCW tail-heaviness index. Panel (d) shows the relationship between the standard deviation of the estimator and coverage probability, stratified by the oracle IPCW tail-heaviness index.

**Detailed results of the clone-censor-weight analysis using real-world medical data**

Fig. S4 shows the patient flow chart. Among 52 672 patients aged ≥18 years who were diagnosed with hemoptysis, 3195 (6.1%) required mechanical ventilation. No patients died on the date of admission; therefore, all 3195 patients were eligible for analysis. The mean (SD) age was 72.3 (14.5) years, and 60.4% of patients were male (n = 1929) (Table S14). The observed 30-d in-hospital all-cause mortality was 35.9% (n = 1147).

Regarding the use of BAE and lung-resection surgery during the grace period (day 0–4), 6 patients (0.2%) underwent both procedures, 602 (18.8%) underwent BAE only, 23 (0.7%) underwent surgery only, and 2564 (80.3%) underwent neither procedure. Among patients who underwent BAE only, corresponding to the intervention protocol, 70 deaths (11.6%) were observed. Among patients who underwent neither procedure, corresponding to the control protocol, 1075 deaths (41.9%) were observed.

Fig. S5 shows the IPCW-weighted Kaplan–Meier curves up to day 30. The weighted survival probability decreased to approximately 0.8 on day 1. Thereafter, the survival curves for the intervention and control strategies gradually diverged, with the intervention strategy consistently showing better survival. In the bootstrap analyses, the weighted mortality was 28.7% (95% CI, 25.9% to 31.2%) under the intervention strategy and 37.5% (35.4% to 39.5%) under the control strategy. The RD was -8.9% (95% CI, -11.8% to -6.1%), and the RR was 0.76 (0.69 to 0.83).

The estimated tail-heaviness index of the IPCW was 12.7 for the intervention strategy and 8.5 for the control strategy. The bootstrap standard errors for the estimated mortality under the intervention and control strategies and for the RD and RR were 0.010, 0.014, 0.014, and 0.036, respectively.

In the exploratory analysis, where pre-existing censoring was not considered, the weighted mortality was 30.6% (95% CI, 27.3% to 33.7%) under the intervention protocol and 39.6% (37.3% to 41.7%) under the control protocol. The RD was -9.0% (95% CI, -12.4% to -5.7%), and the RR was 0.77 (0.69 to 0.85). Patients who were discharged alive were younger than those who remained hospitalized (Fig. S6).

**Figure S4. Patient flow**

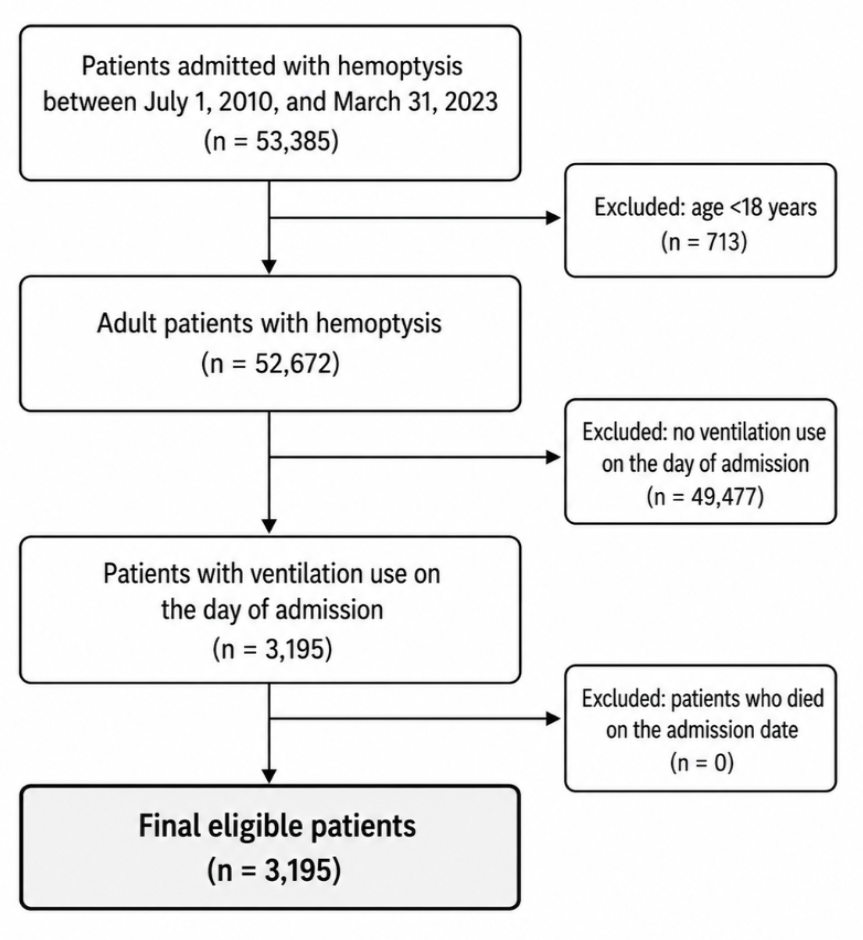

**Table S14. Patient characteristics**

| Total, n (%) | 3195 (100.0) |
| --- | --- |
| Age, mean (SD), years | 71.3 (14.5) |
| Age, median [IQR], years | 74 [66–81] |
| Male, n (%) | 1929 (60.4) |
| Bronchopulmonary carcinoma, n (%) | 171 (5.4) |
| Length of stay, median [IQR], days | 14 [3–29] |
| In-hospital death, n (%) | 1147 (35.9) |
| Day 0 procedures |  |
| BAE, n (%) | 363 (11.4) |
| Surgery, n (%) | 5 (0.2) |
| Bronchoscopy, n (%) | 806 (25.2) |
| Contrast-enhanced CT, n (%) | 1094 (34.2) |
| Mechanical ventilation, n (%) | 3195 (100.0) |
| Intravenous tranexamic acid, n (%) | 1666 (52.1) |
| FFP transfusion, n (%) | 286 (9.0) |
| MAP transfusion, n (%) | 651 (20.4) |
| Platelet transfusion, n (%) | 92 (2.9) |
| Day 0–4 procedures |  |
| BAE, n (%) | 608 (19.0) |
| Surgery, n (%) | 29 (0.9) |
| Bronchoscopy, n (%) | 1201 (37.6) |
| Contrast-enhanced CT, n (%) | 1287 (40.3) |
| Mechanical ventilation, n (%) | 3195 (100.0) |
| Intravenous tranexamic acid, n (%) | 1924 (60.2) |
| FFP transfusion, n (%) | 464 (14.5) |
| MAP transfusion, n (%) | 1123 (35.1) |
| Platelet transfusion, n (%) | 180 (5.6) |

Abbreviations: BAE, bronchial artery embolization; CT, computed tomography; FFP, fresh frozen plasma; IQR, interquartile range; LOS, length of stay; MAP, red blood cell concentrate; SD, standard deviation.

**Figure S5. IPCW-weighted Kaplan–Meier survival curves**

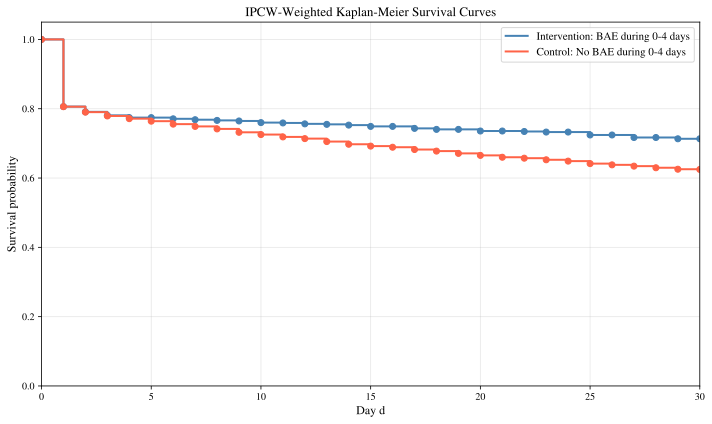

BAE, bronchial artery embolization; IPCW, inverse probability of censoring weight.

**Figure S6. Age distribution according to incident pre-existing censor status**

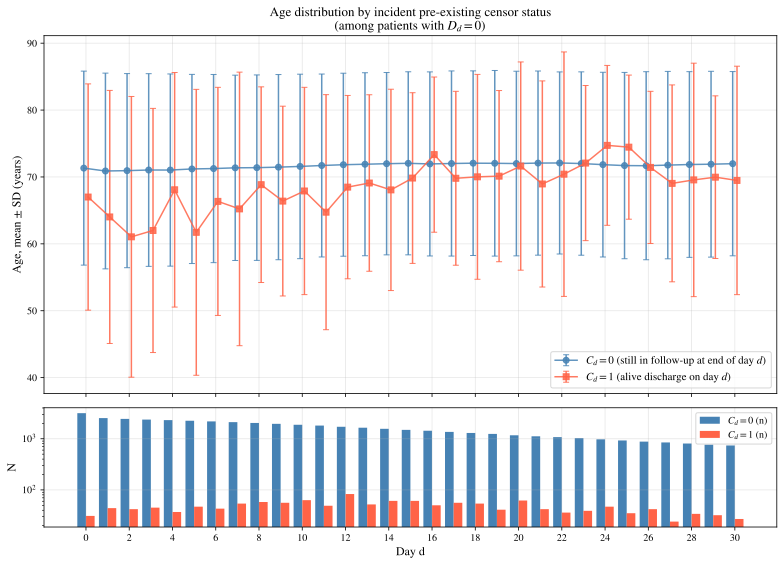

**Explanation for counterintuitive results in Simulation Part 1**

In Simulation Study Part 1, some findings for the intervention strategy in Experiment E with a cutoff of day 0 may appear counterintuitive. Specifically, among clones assigned to the intervention strategy, bias was greater in Experiment E than in Experiment F (0.069 vs 0.026) (Fig. 1). Experiment E also consistently showed substantially higher RMSE than the other experiments, even at a sample size of 10 000, whereas RMSE in the remaining experiments decreased to low levels as the sample size increased (Fig. 2). Coverage probability remained below 0.50 at all sample sizes (Fig. 3).

These findings can be explained as follows. In Experiment E, artificial censoring and pre-existing censoring under the intervention strategy were combined into a single censoring model that included covariates but no terms for time in the intervention strategy (Table 1). Under the intervention strategy, artificial censoring occurs only at the end of the grace period, when all clones who have not yet initiated treatment are censored. The true hazard of the combined censoring process is therefore strongly time dependent.

A model without time terms cannot represent this pattern and instead smooths the sharp cutoff-day increase across the fitted censoring probabilities. When the cutoff is day 0, this ratio is maximized: in an experiment, approximately 90% of clones are censored at time zero, whereas only the small proportion who initiate treatment on day 0 contribute additional person-time. Consequently, the model estimates a constant daily censoring hazard of approximately 30%.

This misspecification affected the analysis in two ways. First, the model failed to apply the large one-time adjustment required for selective treatment initiation on day 0, leaving baseline confounding largely unaddressed. Second, the spuriously elevated daily censoring hazard was compounded multiplicatively over the 30-day follow-up, generating covariate-dependent extreme weights. As a result, the risk sets at later time points were dominated by a small number of clones with extreme weights. Together, these mechanisms produced substantial bias, increased RMSE, and poor confidence-interval coverage.

**Discussion of the real-world medical data analysis results**

As a reference example of how to conduct and report a CCW analysis, as well as to facilitate readers imagine the effect of informative pre-existing censoring, we used real-world data to compare a strategy of performing bronchial artery embolization (BAE) within 5 days of admission with a strategy of withholding it, among patients with severe hemoptysis requiring mechanical ventilation. In a life-threatening disease such as severe hemoptysis, in which deaths occur early after admission, a landmark analysis excludes patients who die or are discharged during the grace period, so its findings apply only to patients who would survive to the landmark and remain under observation. CCW analysis, by contrast, can include every patient who was eligible at admission while keeping eligibility assessment, strategy assignment, and the start of follow-up aligned at the admission date; its findings therefore apply more readily to the population that clinicians face at the time of admission. CCW analysis can also estimate strategy-specific risks for multiple clinically meaningful strategies, provided that practical positivity holds and that enough patients adhere to each strategy—a condition for which the tail-heaviness index serves as one indicator. In our analysis, the estimated tail-heaviness index exceeded 1 and the bootstrap standard errors were relatively small, consistent with the settings that yielded good coverage in our simulations. In addition, the difference in age distribution between patients discharged alive and those who remained hospitalized supported the possibility that discharge alive is an informative censoring event; because patients discharged alive are generally likely to have a better prognosis, censoring them naively may overestimate the in-hospital mortality risk. Because the main purpose of this analysis was to provide a reporting example, however, residual confounding may remain owing to the limited set of covariates, and the results should not be interpreted as definitive evidence on the clinical effectiveness of BAE.

**References.**

1.	Ma X, Wang J. Robust Inference Using Inverse Probability Weighting. *Journal of the American Statistical Association*. Taylor & Francis; 2020 Dec 11;**115**(532):1851–1860.

2.	Yasunaga H. Updated information on the Diagnosis Procedure Combination data. *Ann Clin Epidemiol*. 2024 Oct;**6**(4):106–110.

3.	Yamana H, Moriwaki M, Horiguchi H, Kodan M, Fushimi K, Yasunaga H. Validity of diagnoses, procedures, and laboratory data in Japanese administrative data. *J Epidemiol*. 2017 Oct;**27**(10):476–482.

4.	Kimura Y, Sasabuchi Y, Jo T, et al. Impact of Hemoptysis Etiology and Embolic Agent Type on Prognosis in Patients Undergoing Bronchial Artery Embolization. *Radiol Cardiothorac Imaging*. 2025 June;**7**(3):e240343.
