## Supplementary material for "Pitfalls and Solutions in Clone-Censor-Weight for Target Trial Emulation: Insights from Review, Simulation, and Real-World Analyses": supplemntal file 2

### Supplementary file 2

#### 1 Simulation Experiments

The simulation used longitudinal records indexed by individual $i$ and discrete time $t$. Baseline variables were age, sex, and Charlson comorbidity index (CCI). The time-varying variables were SpO2, treatment $A_t$, death $D_t$, and, in Scenario 3 only, pre-existing censoring $L_t$. Treatment $A_t$ was a one-time event in Scenarios 1–3, and death was absorbing.

The CCW comparison in Scenarios 1–3 used two dynamic protocols with grace-period endpoint $x$. The analyses evaluated $x \in \{0, 2, 4\}$.

- Intervention protocol: before day $x$, the observed treatment process was allowed to operate. At day $x$, patients who had not yet initiated treatment were assigned to initiate treatment. After initiation, no further treatment events were possible because $A_t$ was a one-time treatment event.
- Control protocol: treatment was set to zero for days $t \leq x$. After day $x$, the original treatment process was allowed to operate.

For the CCW analysis, each individual was cloned into each protocol arm. A clone was artificially censored at the first observed time at which its observed treatment history violated the assigned protocol. For the intervention protocol, artificial censoring occurred at day $x$ if treatment had not yet occurred. For the control protocol, artificial censoring occurred at the first treatment event with $t \leq x$.

##### 1.1 Common Baseline Variables

All three scenarios used the same baseline distributions. Age was generated from a two-component normal mixture,

$$\text{AGE}_i \sim \frac{1}{3}N(50, 15^2) + \frac{2}{3}N(80, 10^2).$$

Sex was generated as

$$\Pr(\text{SEX}_i = \text{M}) = \Pr(\text{SEX}_i = \text{F}) = 0.5.$$

CCI took values in $\{0, 1, 2\}$. Its distribution was sex-specific and age-specific. For ages $\leq 50$ ("young") and $\geq 80$ ("old"), the probabilities are shown in Table 1. For ages between 50 and 80, the corresponding young and old probability vectors were linearly interpolated within sex.

##### 1.2 Common SpO2 Process

Each individual had three baseline SpO2-related latent quantities: a pre-COVID SpO2 mean, a COVID-related SpO2 reduction, and a post-COVID target mean. The pre-COVID SpO2 mean was sampled from a beta distribution whose mean and standard deviation were sex-specific and age-specific. The target values are shown in Table 2. For ages between 50 and 80, the corresponding beta shape parameters were linearly interpolated.

Table 1: Sex- and age-specific probabilities used to generate CCI at the young and old age anchors.

| | Pr(CCI = 0) | Pr(CCI = 1) | Pr(CCI = 2) |
|---|---|---|---|
| Young male | 0.4 | 0.3 | 0.3 |
| Old male | 0.1 | 0.3 | 0.6 |
| Young female | 0.5 | 0.4 | 0.1 |
| Old female | 0.2 | 0.4 | 0.4 |

Table 2: Target mean and standard deviation for the pre-COVID SpO2 mean distribution.

| | $\mu$ | $\sigma$ |
|---|---|---|
| Young male | 0.95 | 0.03 |
| Old male | 0.93 | 0.03 |
| Young female | 0.95 | 0.02 |
| Old female | 0.94 | 0.03 |

The acute COVID-related SpO2 reduction was also beta-distributed, with age-specific target mean and standard deviation as shown in Table 3. Again, beta shape parameters were linearly interpolated for ages between 50 and 80. The reduction was truncated so that it could not exceed the pre-COVID SpO2 mean minus 0.01.

Table 3: Target mean and standard deviation for the acute COVID-related SpO2 reduction.

| | $\mu$ | $\sigma$ |
|---|---|---|
| Young | 0.04 | 0.02 |
| Old | 0.06 | 0.02 |

Let $O_i$ denote the pre-COVID SpO2 mean and $E_i$ denote the acute COVID-related SpO2 reduction. The post-COVID target mean was generated as

$$M_i = B_i O_i + (1 - B_i)(O_i - 2E_i), \qquad B_i \sim \text{Beta}(0.06, 0.05),$$

with truncation at 0.01 from below. The observed SpO2 time series was initialized by

$$\text{SpO2}_{i0} = O_i - E_i.$$

If the individual had received treatment, the current target mean was shifted toward the pre-COVID mean:

$$M_{it}^A = \begin{cases} M_i + 0.5(O_i - M_i), & \text{if treatment has occurred before time } t, \\ M_i, & \text{otherwise.} \end{cases}$$

For $t \geq 1$, the transition followed a Gaussian-AR(1) model in the latent space. Let $\phi = 0.5$ and beta precision $\kappa = 500$. The previous SpO2 value was mapped into latent Gaussian space using the beta CDF, an AR(1) Gaussian transition was applied, and the value was mapped back by a beta inverse CDF:

$$U_{i,t-1} = F_{\text{Beta}}\Big(\text{SpO2}_{i,t-1}; \kappa M_{i,t-1}^A, \kappa(1 - M_{i,t-1}^A)\Big),$$

$$Z_{i,t-1} = \Phi^{-1}(U_{i,t-1}), \qquad Z_{it} = \phi Z_{i,t-1} + \sqrt{1 - \phi^2}\varepsilon_{it}, \quad \varepsilon_{it} \sim N(0, 1),$$

$$\text{SpO2}_{it} = F^{-1}_{\text{Beta}}\Big(\Phi(Z_{it}); \kappa M^A_{it}, \kappa(1 - M^A_{it})\Big).$$

After death, subsequent SpO2 values were not generated.

#### 1.3 Normalized Variables Used in Treatment and Outcome Models

The data-generating treatment and death models used normalized age and SpO2 covariates. The pre-existing censoring model in Scenario 3 used normalized age. The age normalizer was

$$A^\star_i = \frac{\text{AGE}_i - \mu_A}{\sigma_A},$$

where $\mu_A$ and $\sigma_A$ denote the mean and standard deviation implied by the age mixture distribution. The capped beta-CDF SpO2 transform was

$$Q_{it} = \min\Big\{F_{\text{Beta}(3.22,0.28)}(\text{SpO2}_{it}),\ 0.5\Big\}.$$

The data-generating treatment and death models used the centered and scaled value

$$S^\star_{it} = 8(Q_{it} - 0.25).$$

In the analysis models below, current normalized SpO2 refers to $Q_{it}$ and baseline normalized SpO2 refers to $Q_{i0}$.

Let $I^M_i = \mathbf{1}\{\text{SEX}_i = \text{M}\}$ and let $H^A_{it}$ be the cumulative treatment history available for the transition from $t$ to $t + 1$.

#### 1.4 Data-Generating Scenarios

**Scenario 1: time-varying confounding without pre-existing censoring.** Scenario 1 included time-varying confounding by SpO2: current SpO2 affected treatment initiation, and prior SpO2 affected subsequent death. Treatment could occur at most once. Among individuals not already treated and not dead, treatment was generated by

$$\operatorname{logit}\Pr(A_{it} = 1 \mid \mathcal{H}_{it}) = -4.0 - 0.05 I^M_i + 0.25 A^\star_i + 0.25\text{CCI}_i - 1.0 S^\star_{it} + \exp(-0.1t).$$

Death was generated as an absorbing event. For the transition from $t$ to $t + 1$ among those alive,

$$\operatorname{logit}\Pr(D_{i,t+1} = 1 \mid D_{it} = 0, \mathcal{H}_{it}) = -5.0 + 0.8 I^M_i + 0.8 A^\star_i + 0.6\text{CCI}_i - 3.0 S^\star_{it} - 2.0 H^A_{it}.$$

**Scenario 2: no time-varying confounding in treatment assignment.** Scenario 2 kept the same baseline-variable, SpO2, and death processes as Scenario 1, but removed SpO2 from the treatment initiation model. Thus, treatment initiation depended on baseline variables but not on the current time-varying SpO2 value:

$$\operatorname{logit}\Pr(A_{it} = 1 \mid \mathcal{H}_{it}) = -4.0 - 0.05 I^M_i + 0.25 A^\star_i + 0.25\text{CCI}_i + \exp(-0.1t).$$

The death model was the same as in Scenario 1. SpO2 remained prognostic for death, but it was not a cause of treatment initiation in this scenario.

**Scenario 3: time-varying confounding with pre-existing informative censoring.** Scenario 3 used the Scenario 1 structure with an additional pre-existing censoring process $L_t$. The same treatment model and the death model were the same as in Scenario 1.

The pre-existing censoring process was absorbing, with no censoring at time zero. Among individuals not already censored,

$$\operatorname{logit}\Pr(L_{it} = 1 \mid \mathcal{H}_{it}) = -3.5 - 0.3 I_i^M - 0.8 A_i^\star - 0.6\mathrm{CCI}_i.$$

This censoring process depended only on baseline covariates, independently.

#### 1.5 Parameter Sets for Part 2

Part 2 used Experiment A. Let us denote the reference setting as `a1d1`. Two one-dimensional parameter families were considered, with $x \in \{0.5, 1, 2, 4, 8\}$. In the `axd1` family, the treatment-assignment covariate coefficients were multiplied by $x$, giving

$$(\beta_M^A, \beta_{\mathrm{age}}^A, \beta_{\mathrm{CCI}}^A, \beta_{\mathrm{SpO2}}^A) = x(-0.05, 0.25, 0.25, -1.0),$$

while the death model was kept at the `a1d1` values. In the `a1dx` family, the treatment model was kept at the `a1d1` values and the death-model prognostic coefficients were multiplied by $x$:

$$(\beta_M^D, \beta_{\mathrm{age}}^D, \beta_{\mathrm{CCI}}^D, \beta_{\mathrm{SpO2}}^D) = x(0.8, 0.8, 0.6, -3.0).$$

The treatment-effect coefficient in the death model remained $-2.0$, and all other simulation and analysis settings were unchanged.

#### 1.6 Analysis Experiments

The experiments used three IPCW modeling approaches. In the first approach, a single pooled logistic model was fitted for the combined censoring event. When no pre-existing censoring was present, the combined event was artificial censoring only. When pre-existing censoring was present, the combined event was the union of artificial and pre-existing censoring. In the second approach, separate pooled logistic models were fitted for artificial censoring and pre-existing censoring, and the final IPCW was the product of the component weights. In the third approach, only artificial censoring was modeled, and pre-existing censoring was not adjusted for in the IPCW model. The experiment configurations are summarized in Table 4.

Table 4: Simulation experiments and IPCW denominator models. The table shows the linear predictors used in the pooled logistic denominator models for censoring. Here $Q_{it}$ denotes current normalized SpO2, $Q_{i0}$ denotes baseline normalized SpO2, $I_i^M$ denotes male sex, and $\delta_t$ denotes a categorical time effect. Coefficients are nuisance parameters estimated from the simulated data.

| Experiment | Scenario | IPCW modeling approach | Denominator models |
|---|---|---|---|
| A | Scenario 1 | Artificial-censoring model only | Control arm: $\operatorname{logit}\Pr(C_{it}^{\text{art}}=1 \mid \cdot) = \eta_0+\delta_t+\eta_1\text{AGE}_i+\eta_2 I_i^M+\eta_3\text{CCI}_i+\eta_4 Q_{it}$. Intervention arm: $\operatorname{logit}\Pr(C_{it}^{\text{art}}=1 \mid \cdot) = \eta_0+\eta_1\text{AGE}_i+\eta_2 I_i^M+\eta_3\text{CCI}_i+\eta_4 Q_{it}$. |
| B | Scenario 1 | Artificial-censoring model only | Control arm: $\operatorname{logit}\Pr(C_{it}^{\text{art}}=1 \mid \cdot) = \eta_0+\delta_t+\eta_1\text{AGE}_i+\eta_2 I_i^M+\eta_3\text{CCI}_i+\eta_4 Q_{i0}$. Intervention arm: $\operatorname{logit}\Pr(C_{it}^{\text{art}}=1 \mid \cdot) = \eta_0+\eta_1\text{AGE}_i+\eta_2 I_i^M+\eta_3\text{CCI}_i+\eta_4 Q_{i0}$. |
| C | Scenario 2 | Artificial-censoring model only | Control arm: $\operatorname{logit}\Pr(C_{it}^{\text{art}}=1 \mid \cdot) = \eta_0+\delta_t+\eta_1\text{AGE}_i+\eta_2 I_i^M+\eta_3\text{CCI}_i+\eta_4 Q_{i0}$. Intervention arm: $\operatorname{logit}\Pr(C_{it}^{\text{art}}=1 \mid \cdot) = \eta_0+\eta_1\text{AGE}_i+\eta_2 I_i^M+\eta_3\text{CCI}_i+\eta_4 Q_{i0}$. |
| D | Scenario 3 | Separate models for artificial and pre-existing censoring | Artificial-censoring model, control arm: $\operatorname{logit}\Pr(C_{it}^{\text{art}}=1 \mid \cdot) = \eta_0+\delta_t+\eta_1\text{AGE}_i+\eta_2 I_i^M+\eta_3\text{CCI}_i+\eta_4 Q_{it}$. Artificial-censoring model, intervention arm: $\operatorname{logit}\Pr(C_{it}^{\text{art}}=1 \mid \cdot) = \eta_0+\eta_1\text{AGE}_i+\eta_2 I_i^M+\eta_3\text{CCI}_i+\eta_4 Q_{it}$. Pre-existing censoring model, both arms: $\operatorname{logit}\Pr(L_{it}=1 \mid \cdot) = \zeta_0+\zeta_1\text{AGE}_i+\zeta_2 I_i^M+\zeta_3\text{CCI}_i$. |
| E | Scenario 3 | Joint model for artificial and pre-existing censoring | Let $C_{it}^{\text{all}}=1$ denote either artificial censoring or pre-existing censoring at time $t$. Control arm: $\operatorname{logit}\Pr(C_{it}^{\text{all}}=1 \mid \cdot) = \eta_0+\eta_1\text{AGE}_i+\eta_2 I_i^M+\eta_3\text{CCI}_i+\eta_4 Q_{it}$. Intervention arm: $\operatorname{logit}\Pr(C_{it}^{\text{all}}=1 \mid \cdot) = \eta_0+\eta_1\text{AGE}_i+\eta_2 I_i^M+\eta_3\text{CCI}_i+\eta_4 Q_{it}$. |

Continued on next page

Table 4: Simulation experiments and IPCW denominator models (continued).

| Experiment | Scenario | IPCW modeling approach | Denominator models |
|---|---|---|---|
| F | Scenario 3 | Artificial-censoring model only | Control arm: $\operatorname{logit}\Pr(C_{it}^{\mathrm{art}}=1 \mid \cdot)=\eta_0+\delta_t+\eta_1\mathrm{AGE}_i+\eta_2 I_i^M+\eta_3\mathrm{CCI}_i+\eta_4 Q_{it}$. Intervention arm: $\operatorname{logit}\Pr(C_{it}^{\mathrm{art}}=1 \mid \cdot)=\eta_0+\eta_1\mathrm{AGE}_i+\eta_2 I_i^M+\eta_3\mathrm{CCI}_i+\eta_4 Q_{it}$. |

### 2 Theoretical Justification of the IPCW Tail-Index Diagnostics

Fix a protocol arm $a$ and an interval endpoint $t+1$. This section establishes which inverse-probability factors determine the second moment of the IPCW hazard score and thereby justifies the two tail indices used in the analysis.

**Observed-data structure and assumptions.** Let $V_i$ contain baseline variables, let $X_{is}$ contain the covariate and past-treatment information available immediately before the time-$s$ adherence or censoring decision, let $D_{is}$ be the absorbing death indicator, and let $C_{is}^a$ be the protocol-specific censoring or deviation indicator. The within-interval ordering is

$$D_{is} \longrightarrow X_{is} \longrightarrow C_{is}^a \longrightarrow D_{i,s+1}.$$

Thus, for the endpoint $t+1$, the arm-specific observed trajectory is

$$\mathcal{O}_i^a = \big(V_i, D_{i0}, X_{i0}, C_{i0}^a, D_{i1}, X_{i1}, C_{i1}^a, \ldots, D_{it}, X_{it}, C_{it}^a, D_{i,t+1}\big),$$

where $D_{i,s+1}$ is observed only while the individual is alive and uncensored through time $s$. The history immediately before $C_{is}^a$ is

$$\mathcal{H}_{is}^a = \sigma\big(V_i, D_{i,0:s}, X_{i,0:s}, C_{i,0:s-1}^a\big).$$

Write $\bar{C}_{i,s}^a = (C_{i0}^a, \ldots, C_{is}^a)$, with $\bar{C}_{i,-1}^a = 0$, and, for $s = 0, \ldots, t$, define

$$E_{is}^a = \mathbf{1}\{D_{is}=0,\ \bar{C}_{i,s-1}^a = 0\}, \qquad G_{is}^a = \Pr(C_{is}^a = 0 \mid \mathcal{H}_{is}^a, E_{is}^a = 1).$$

Here $E_{is}^a$ is the pre-censoring risk indicator. Values of $G_{is}^a$ outside this risk set may be set to one. Define

$$H_{i,t-1}^a = \prod_{s=0}^{t-1} G_{is}^a, \qquad R_{it}^a = E_{it}^a \mathbf{1}(C_{it}^a = 0), \qquad W_{it}^a = \frac{1}{H_{i,t-1}^a G_{it}^a},$$

with $H_{i,-1}^a = 1$. The counterfactual death trajectory under protocol $a$ with protocol-specific censoring eliminated is

$$\bar{D}_{i,0:t+1}^a = (D_{i0}^a, D_{i1}^a, \ldots, D_{i,t+1}^a),$$

and the target hazard is

$$p_{t+1}^a = \Pr(D_{i,t+1}^a = 1 \mid D_{it}^a = 0).$$

The following conditions are assumed, with time-indexed statements applying for $s = 0, \ldots, t$.

(A1) The trajectories $\mathcal{O}_1^a, \ldots, \mathcal{O}_n^a$ are i.i.d.; protocol $a$ is well defined; there is no interference; and $D_{iu}^a \in \{0,1\}$ with $D_{iu}^a \le D_{i,u+1}^a$ for $u = 0, \ldots, t$.

(A2) Consistency holds:

$$D_{iu} = D_{iu}^a \quad \text{on } \{\bar{C}_{i,u-1}^a = 0\}, \qquad u = 0, \ldots, t+1.$$

(A3) Censoring is sequentially exchangeable among eligible individuals:

$$C_{is}^a \perp\!\!\!\perp (D_{is}^a, D_{i,s+1}^a, \ldots, D_{i,t+1}^a) \mid \mathcal{H}_{is}^a, E_{is}^a = 1.$$

(A4) Weak positivity holds:

$$\Pr(E_{is}^a = 1) > 0, \qquad \Pr(G_{is}^a > 0 \mid E_{is}^a = 1) = 1.$$

No uniform lower bound on $G_{is}^a$ is imposed.

(A5) The counterfactual risk set is nonempty: $\Pr(D_{it}^a = 0) > 0$.

For estimation, let $G_{is}^a(\gamma) = G_s^a(\mathcal{H}_{is}^a; \gamma)$ be a model for $G_{is}^a$, and let $\hat{\gamma}$ be its fitted parameter. Separately from (A1)–(A5), assume correct specification: there is a $\gamma_0$ such that

$$G_{is}^a(\gamma_0) = G_{is}^a \quad \text{a.s. on } \{E_{is}^a = 1\}, \qquad s = 0, \ldots, t. \tag{M1}$$

With

$$\psi_{it}^a(p, \gamma) = W_{it}^a(\gamma) R_{it}^a (D_{i,t+1} - p), \qquad W_{it}^a(\gamma) = \prod_{s=0}^{t} \{G_{is}^a(\gamma)\}^{-1},$$

the equation $\mathbb{P}_n\{\psi_{it}^a(p, \hat{\gamma})\} = 0$ gives

$$\hat{p}_{t+1}^a = \frac{\sum_{i=1}^n W_{it}^a(\hat{\gamma}) R_{it}^a D_{i,t+1}}{\sum_{i=1}^n W_{it}^a(\hat{\gamma}) R_{it}^a},$$

when the denominator is positive. Hence $\hat{S}_u^a = \prod_{r=0}^{u-1}(1 - \hat{p}_{r+1}^a)$ is the IPCW Kaplan–Meier estimator.

**Proposition 1 (unbiasedness and variance characterization).** Under (A1)–(A5) and (M1),

$$\mathbb{E}\{\psi_{it}^a(p_{t+1}^a, \gamma_0)\} = 0.$$

If $0 < p_{t+1}^a < 1$, then

$$\operatorname{Var}\{\psi_{it}^a(p_{t+1}^a, \gamma_0)\} < \infty \quad \iff \quad \mathbb{E}\left\{ \frac{E_{it}^a}{(H_{i,t-1}^a)^2 G_{it}^a} \right\} < \infty. \tag{1}$$

*Proof.* Set

$$K_{is}^a = \prod_{r=0}^{s} \frac{\mathbf{1}(C_{ir}^a = 0)}{G_{ir}^a}, \qquad K_{i,-1}^a = 1, \qquad U_{it}^a = \mathbf{1}(D_{it}^a = 0)(D_{i,t+1}^a - p_{t+1}^a).$$

Consistency and (M1) imply $\psi_{it}^a(p_{t+1}^a, \gamma_0) = K_{it}^a U_{it}^a$. Because death is absorbing, for every $s \le t$, $K_{i,s-1}^a U_{it}^a = K_{i,s-1}^a E_{is}^a U_{it}^a$. Sequential exchangeability and the definition of $G_{is}^a$ therefore give

$$\begin{aligned} \mathbb{E}(K_{is}^a U_{it}^a) &= \mathbb{E}\left[ K_{i,s-1}^a E_{is}^a \mathbb{E}\left\{ \frac{\mathbf{1}(C_{is}^a = 0)}{G_{is}^a} U_{it}^a \;\middle|\; \mathcal{H}_{is}^a, E_{is}^a = 1 \right\} \right] \\ &= \mathbb{E}\Big[ K_{i,s-1}^a E_{is}^a \mathbb{E}(U_{it}^a \mid \mathcal{H}_{is}^a, E_{is}^a = 1) \Big] = \mathbb{E}(K_{i,s-1}^a U_{it}^a). \end{aligned}$$

Applying the same peeling identity first to $|U^a_{it}|$ establishes integrability, since $\mathbb{E}|K^a_{it}U^a_{it}| = \mathbb{E}|U^a_{it}| \leq 1$. Peeling this identity backward from $s = t$ to $s = 0$ yields

$$\mathbb{E}\{\psi^a_{it}(p^a_{t+1}, \gamma_0)\} = \mathbb{E}(U^a_{it}) = \Pr(D^a_{it} = 0)\{\mathbb{E}(D^a_{i,t+1} \mid D^a_{it} = 0) - p^a_{t+1}\} = 0.$$

For the second moment, define

$$q^a_t(\mathcal{H}^a_{it}) = \mathbb{E}\Big[(D^a_{i,t+1} - p^a_{t+1})^2 \mid \mathcal{H}^a_{it}, E^a_{it} = 1\Big].$$

Conditioning the squared score on the pre-censoring history, then applying consistency and sequential exchangeability at time $t$, gives

$$\mathbb{E}\Big[\psi^a_{it}(p^a_{t+1}, \gamma_0)^2 \mid \mathcal{H}^a_{it}\Big] = \frac{E^a_{it}}{(H^a_{i,t-1})^2 G^a_{it}} q^a_t(\mathcal{H}^a_{it}). \tag{2}$$

The single power of $(G^a_{it})^{-1}$ in (2) results from integrating out $\mathbf{1}(C^a_{it} = 0)$; the variance driver is therefore

$$V^a_{it} = \frac{1}{(H^a_{i,t-1})^2 G^a_{it}}.$$

For binary $D^a_{i,t+1}$ and $0 < p^a_{t+1} < 1$,

$$0 < c^a_t := \min\{(p^a_{t+1})^2, (1 - p^a_{t+1})^2\} \leq q^a_t(\mathcal{H}^a_{it}) \leq 1.$$

Taking expectations in (2) consequently gives

$$c^a_t \mathbb{E}(E^a_{it}V^a_{it}) \leq \mathrm{Var}\{\psi^a_{it}(p^a_{t+1}, \gamma_0)\} \leq \mathbb{E}(E^a_{it}V^a_{it}),$$

which proves (1). □

Hereafter, *infinite variance* refers specifically to the infinite second moment of this untruncated IPCW hazard score, which is the object assessed by the diagnostics. It does not refer to the finite-sample variance of the bounded ratio $\hat{p}^a_{t+1}$.

**Proposition 2 (variance decomposition underlying the two diagnostics).** Let

$$X^a_{H,it} = (H^a_{i,t-1})^{-2}, \qquad X^a_{G,it} = (G^a_{it})^{-1}, \qquad \mu^a_{H,t} = \mathbb{E}(E^a_{it}X^a_{H,it}).$$

Because $X^a_{G,it} \geq 1$, $\mu^a_{H,t} = \infty$ implies $\mathbb{E}(E^a_{it}V^a_{it}) = \infty$. If $\mu^a_{H,t} < \infty$, define the probability measure $Q^a_t$ by

$$\frac{\mathrm{d}Q^a_t}{\mathrm{d}P} = \frac{E^a_{it}X^a_{H,it}}{\mu^a_{H,t}}.$$

Then the variance driver has the exact factorization

$$\mathbb{E}(E^a_{it}V^a_{it}) = \mu^a_{H,t}\, \mathbb{E}_{Q^a_t}(X^a_{G,it}). \tag{3}$$

Thus, when $\mu^a_{H,t} < \infty$, the score variance is finite if and only if $\mathbb{E}_{Q^a_t}(X^a_{G,it}) < \infty$. Equation (3) follows directly from $V^a_{it} = X^a_{H,it}X^a_{G,it}$ and the definition of $Q^a_t$; it requires no independence between the historical and current probabilities. □

At $t = 0$, $X^a_{H,i0} = 1$ by definition, so its first moment is finite and no historical tail index is required.

**Tail-index diagnostics.** Let $P_t^a = P(\cdot \mid E_{it}^a = 1)$. For each row in the pre-censoring risk set $\{E_{it}^a = 1\}$, let $g_j^a$ and $h_j^a$ be the fitted values of $G_{it}^a$ and $H_{i,t-1}^a$. The sample tail variables and analysis weights corresponding to $P_t^a$ and $Q_t^a$ are

$$(Z_j, w_j) = \begin{cases} ((h_j^a)^{-2}, 1), & \text{historical probability accumulation,} \\ ((g_j^a)^{-1}, (h_j^a)^{-2}), & \text{current uncensoring probability.} \end{cases} \tag{4}$$

In particular, the historical rows are selected before observing $C_{it}^a$; restricting them to $C_{it}^a = 0$ would change the target law $P_t^a$ unless inverse-$G_{it}^a$ reweighting or an additional tail-equivalence condition were imposed.

The Pareto-type assumption is needed only to translate these two moment targets into tail-index thresholds. Specifically, suppose the historical factor under $P_t^a$ has a regularly varying upper tail

$$P_t^a(X_{H,it}^a > z) \sim z^{-\alpha_{H,t}^a} L_H(z), \tag{5}$$

and, whenever $\mu_{H,t}^a < \infty$, the current factor under $Q_t^a$ satisfies

$$Q_t^a(X_{G,it}^a > z) \sim z^{-\alpha_{G,t}^a} L_G(z), \tag{6}$$

where $\alpha_{H,t}^a, \alpha_{G,t}^a > 0$ and $L_H, L_G$ are slowly varying. Since $\mathbb{E}(X) = \int_0^\infty \Pr(X > z)\,\mathrm{d}z$ for non-negative $X$, such a first moment is finite when $\alpha > 1$ and infinite when $\alpha < 1$. Propositions 1–2 therefore give

$$\begin{aligned} \alpha_{H,t}^a < 1 \quad &\Longrightarrow \quad \operatorname{Var}\{\psi_{it}^a(p_{t+1}^a, \gamma_0)\} = \infty, \\ \alpha_{H,t}^a > 1,\ \alpha_{G,t}^a < 1 \quad &\Longrightarrow \quad \operatorname{Var}\{\psi_{it}^a(p_{t+1}^a, \gamma_0)\} = \infty, \\ \alpha_{H,t}^a > 1,\ \alpha_{G,t}^a > 1 \quad &\Longrightarrow \quad \operatorname{Var}\{\psi_{it}^a(p_{t+1}^a, \gamma_0)\} < \infty. \end{aligned} \tag{7}$$

At $\alpha = 1$, finiteness depends on the slowly varying term, so the index alone is inconclusive. Thus regular variation is a calibration assumption for the Pareto indices and their threshold at 1; it is not required for the exact variance characterization in Propositions 1–2.

Order the positive finite pairs so that $Z_{(1)} \geq \cdots \geq Z_{(m)}$. For $k < m$, the weighted Hill estimator and the corresponding Pareto tail-index estimator are

$$\hat{\gamma}_k = \frac{\sum_{j=1}^k w_{(j)} \log\{Z_{(j)}/Z_{(k+1)}\}}{\sum_{j=1}^k w_{(j)}}, \qquad \hat{\alpha}_k = \frac{1}{\hat{\gamma}_k}. \tag{8}$$

Under (5)–(6), consistent estimation of the uncensoring probabilities, an intermediate sequence of tail sizes, and the usual regularity conditions for the unweighted or weighted Hill estimator (including negligible plug-in error at the relevant upper order statistics), (8) targets the corresponding population tail index. A population index below 1 implies an infinite variance-relevant first moment; an estimate below 1 is evidence of that behavior. An estimate above 1 supports finiteness only when the other factor in (3) is also finite, and estimates near 1 are necessarily inconclusive. The current index has the population interpretation in (6) only after the historical first moment is finite.
