## supplemental file 4 for "Pitfalls and Solutions in Clone-Censor-Weight for Target Trial Emulation: Insights from Review, Simulation, and Real-World Analyses"

**Checklist for Clone–Censor–Weight Analysis**

**1. Design**

**1-1. Eligibility Criteria and Treatment Strategies**

- Have the eligibility criteria and treatment strategies been clearly defined?
  - A sufficient number of eligible patients should be expected to adhere to each strategy.
  - For a point treatment, the basic comparison is generally between “initiate treatment during the grace period” and “do not initiate treatment during the grace period.” Additional conditions, such as not initiating another treatment during the grace period, should be explicitly incorporated into the strategy when necessary.
  - For a sustained treatment, several strategies may be considered. Treatment use after initiation should therefore be explicitly defined. Examples include:
    - Initiate treatment during the grace period.
    - Initiate treatment during the grace period and continue it thereafter.
    - Do not initiate treatment during the grace period and remain untreated thereafter.
    - Do not initiate treatment during the grace period, but allow initiation after the grace period.
  - Is the grace period clinically reasonable, while still allowing a sufficient number of patients to adhere to each strategy?

**1-2. Pre-existing Censoring**

- Are there any forms of pre-existing censoring, such as loss to follow-up, discharge alive, or loss of insurance eligibility, and are they likely to be informative?
  - If so, has it been determined whether IPCW will be used to estimate outcomes under a hypothetical setting in which pre-existing censoring does not occur? This will generally be the preferred approach.

**1-3. Competing Events**

- If the outcome is subject to competing events, has an approach appropriate to the research objective been selected^1^?
  - Treatment policy
  - Composite
  - While-on-treatment
  - Hypothetical
  - Principal stratum
- If a hypothetical strategy is selected, can the estimates under a setting in which the competing event does not occur be interpreted causally and clinically?
  - If such an interpretation is difficult, another estimand strategy should be considered.

**2. Implementation**

**2-1. Data Preparation**

- Have the data for each patient been transformed into an appropriate time-split format, such as daily, weekly, or monthly records?
- Have clones corresponding to the number of treatment strategies been created for each patient and assigned to the respective strategies?
- Has artificial censoring been correctly applied when each clone deviates from its assigned strategy?
- Has the order of events been prespecified when the outcome, artificial censoring, pre-existing censoring, or a competing event occurs at the same time?
  - An unclear event-ordering rule may lead to incorrect definition of the risk sets used in the IPCW models.

**2-2. IPCW Models**

- Has a separate IPCW model for artificial censoring been constructed for each strategy?
  - Because the mechanism of artificial censoring differs across strategies, separate models are generally required.
- Does each IPCW model include only records belonging to the risk set in which the relevant censoring event can occur?
  - Records for which the censoring indicator must always equal 0 should not be included.
  - For example, under a strategy defined as “initiate treatment during days 0–4,” artificial censoring can occur only on day 4. Therefore, only records corresponding to day 4 should be included in that artificial-censoring model.
  - Records after the occurrence of an outcome or censoring event should be excluded from the risk set.
  - Incorrect risk-set specification can constitute a major implementation error in IPCW analysis.
- If the probability of censoring varies over follow-up, has a time variable been incorporated into the model using an appropriate form, such as categorical indicators or splines?
  - A time variable is unnecessary when censoring can occur at only one specific time point, although including one is unlikely to be harmful if the risk set is correctly defined.
- When adjusting for informative pre-existing censoring or competing events, does the modeling approach reflect the distinct mechanisms underlying artificial censoring, pre-existing censoring, and competing events?

**Estimation and Diagnostics**

- Has the final IPCW been applied using a weighted Kaplan–Meier estimator or another weighted outcome model appropriate for the target estimand?
- Have confidence intervals been calculated using a method that accounts for correlations between clones originating from the same patient, such as patient-level bootstrap resampling?
- Has the tail-heaviness index been calculated separately for each strategy?
- If the tail-heaviness index is below 1, has the high likelihood that conventional asymptotic inference and bootstrap confidence intervals may be invalid been addressed?
  - First, check for model misspecification and data-processing errors.
  - If no such problems are identified, reconsider the eligibility criteria, strategy definition, or grace period within clinically reasonable limits.
- Even when the tail-heaviness index is 1 or greater, has the bootstrap standard error been evaluated as an additional diagnostic?
  - A large standard error may indicate unstable finite-sample inference.
  - Values such as 0.01, 0.05, and 0.10 should not be interpreted as universal cutoffs.

Note that this checklist is not intended to be comprehensive but instead highlights key aspects of clone–censor–weight analysis that investigators may misunderstand.

**References.**

1.	Kahan BC, Hindley J, Edwards M, Cro S, Morris TP. The estimands framework: a primer on the ICH E9(R1) addendum. *BMJ*. Clinical research ed.; 2024 Jan;**384**:e076316.
